# Contribution of nosocomial transmission to *Klebsiella pneumoniae* neonatal sepsis in Africa and South Asia: analysis of infection clusters inferred from pathogen genomics and temporal data

**DOI:** 10.1101/2025.11.15.25340095

**Authors:** Erkison Ewomazino Odih, Jabir A Abdulahi, Anne V Amulele, Matthew Bates, Eva Heinz, Weiming Hu, Kajal Jain, Rindidzani Magobo, Courtney P Olwagen, John M Tembo, Tolbert Sonda, Jonathan Strysko, Caroline C Tigoi, Kyle Bittinger, Jennifer Cornick, Ebenezer Foster-Nyarko, Wilson Gumbi, Steven M Jones, Chileshe L Musyani, Carolyn M McGann, Ahmed M Moustafa, Patrick Musicha, James CL Mwansa, Moreka L Ndumba, Thomas D Stanton, Donwilliams O Omuoyo, Oliver Pearse, Laura T Phillips, Paul J Planet, Charlene MC Rodrigues, Fatou Secka, Kirsty Sands, Erin Theiller, Allan M Zuza, Sulagna Basu, Grace J Chan, Kenneth C Iregbu, Jean-Baptiste Mazarati, Semaria Solomon Alemayehu, Timothy R Walsh, Rabaab Zahra, Angela Dramowski, Sombo Fwoloshi, Appiah-Korang Labi, Lola Madrid, Noah Obeng-Nkrumah, David Ojok, Boaz D Wadugu, Andrew C Whitelaw, Anudita Bhargava, Atul Jindal, Ramesh K Agarwal, Alexander M Aiken, James A Berkley, Susan E Coffin, Nicholas A Feasey, Nelesh P Govender, Davidson H Hamer, Shabir A Madhi, M Jeeva Sankar, Kelly L Wyres, Kathryn E Holt

## Abstract

**Background:** *Klebsiella pneumoniae* is the leading cause of sepsis among neonates in low- and middle-income countries (LMICs) in Africa and Asia, contributing substantially to the overall burden of antimicrobial resistant (AMR) infections and mortality among neonates globally. Pathogen sequencing has been used to investigate case clusters and confirm nosocomial transmission in a small number of neonatal units. Here we utilise pathogen sequence data to estimate the fraction of *K. pneumoniae* neonatal sepsis attributable to nosocomial transmission in African and South Asian countries.

**Methods and Findings:** We estimated the proportion of invasive *K. pneumoniae* disease involved in nosocomial transmission clusters in a given neonatal unit, using single-linkage clustering based on pairwise temporal and genetic distances estimated from bacterial whole-genome sequences aggregated from 10 contributing studies. Analysing 1,523 *K. pneumoniae* isolates from 27 units in 13 countries in Africa and South Asia between 2013 and 2023, we inferred 156 nosocomial transmission clusters, ranging from 2 to 188 neonates each (83 of the clusters comprised ≥3 cases). Overall, we estimated that 1,035 neonatal infections (68.0%) were part of nosocomial transmission clusters. Excluding the first infection in each cluster as a potential index case, we estimate at least 879 (57.7%) infections were acquired via nosocomial transmission. Sensitivity analyses showed that results were robust to the choice of genetic distance estimation methods and thresholds used to define clusters, and cluster estimates were stable over temporal distance thresholds ranging from 2 to 8 weeks. Isolates were mostly extended-spectrum beta-lactamase (ESBL) producers (90.9%) and included 172 multi-locus sequence types (STs). Fourteen STs, including several globally recognised multidrug-resistant lineages, were associated with transmission clusters at multiple units and these were collectively responsible for two-thirds of all infections. Carriage of carbapenemase genes (adjusted odds ratio, aOR = 2.08 [95% confidence interval, CI: 1.04–4.14], p=0.02) and ESBL genes (aOR = 2.48 [95% CI: 1.26–4.90] p=0.006) were significantly positively associated with transmission.

**Conclusions:** Nosocomial transmission contributes to a substantial proportion of *K. pneumoniae* sepsis in neonatal care units in Africa and South Asia. Reducing transmission within these settings through improved infection prevention and control and other measures could substantially reduce the neonatal sepsis burden. A high burden of transmission clusters is associated with the same drug-resistant lineages that are recognised as high-risk clones associated with hospital outbreaks in high-income countries, indicating global connectivity of the AMR pathogen population.

**Author Summary:** *Why Was This Study Done?:*

- *Klebsiella pneumoniae* is the leading cause of sepsis among neonates in low- and middle-income countries (LMICs) in Africa and Asia, and the infections are difficult to treat due to rising rates of antimicrobial resistance.
- Invasive bacterial diseases are typically transmitted to neonates from their mothers before, during or soon after birth (vertical transmission) or from the hospital environment and healthcare workers (horizontal transmission).
- The fraction of *K. pneumoniae* neonatal sepsis cases attributable to horizontal transmission is unknown, but this information is important to understand the role of infection prevention and control (IPC) measures in lowering disease burden.

*What Did the Researchers Do and Find?:*

- We developed a simple method to detect transmission clusters from genetic and temporal distance data and found the method to be robust to the choice of genetic and temporal distance thresholds.
- We applied this method to detect transmission clusters among 1,523 *K. pneumoniae* neonatal sepsis cases from 10 studies and 27 hospitals across Africa and South Asia.
- We estimate over half of sepsis cases (68.0%) were part of a transmission cluster, and by excluding the hypothetical index case for each cluster we estimate at least 57.7% of infections were acquired via nosocomial transmission.
- Most of the isolates (90.9%) were extended-spectrum beta-lactamase (ESBL) producers (conferring resistance to third-generation cephalosporin antibiotics), and carriage of ESBL and carbapenemase genes (conferring resistance to carbapenem antibiotics) were positively associated with transmission.
- Fourteen genetic lineages were associated with clusters in multiple neonatal units, together accounting for two-thirds of all infections.
- Many of these same lineages are common causes of drug-resistant hospital outbreaks in high-income countries.

*What Do These Findings Mean?:*

- A substantial proportion of *K. pneumoniae* neonatal sepsis cases are potentially preventable with improvements in IPC in neonatal units.
- Further work is needed to identify and better understand transmission routes and risk factors for transmission to support the implementation of effective and scalable IPC solutions.
- Our findings highlight the importance of genomic surveillance to support IPC interventions for *K. pneumoniae* and other pathogens, and reveal many of the same ‘drug-resistant problem clones’ are responsible for hospital outbreaks across high-and low-income countries.
- The high rates of ESBL gene carriage among isolates in this study indicates that empirical treatment based on the current WHO guidelines may result in high rates of treatment failure.
- The limitations of this study include the lack of sufficient clinical data to allow high-resolution investigation of transmission dynamics, as well as facility-level data to investigate contributors to the observed differences in transmission burden across sites.

## Introduction

Sepsis is a leading cause of death among neonates, particularly in Africa and Asia, where the highest global incidence of neonatal mortality was recorded in 2022, with over 21 deaths per 1000 live births [1–3]. Bacterial infections are the most common cause of neonatal sepsis globally, with the predominant pathogens varying across geographical regions [4–10]. In Africa and Asia, *K. pneumoniae* is the primary cause of neonatal sepsis, and displays high rates of resistance to empirical antimicrobials recommended by WHO [5,11]. This makes the infections difficult to treat which contributes to higher neonatal mortality rates, mostly in low-and middle-income countries (LMICs) where the disease burden is highest and access to high-quality medical care and effective antibiotics is often limited [12,13].

Invasive bacterial disease in neonates is associated with two main transmission routes; vertical transmission from the mother, and horizontal transmission from other sources including the hospital environment and healthcare workers [8,14,15]. As such, efforts to reduce the burden of neonatal sepsis focus on intrapartum antimicrobial prophylaxis [16], improving infection prevention and control (IPC) practices, early case identification, and sepsis management [17].

*K. pneumoniae* is well-established as a leading cause of nosocomial infection outbreaks in hospitals globally [18], more likely to cause clusters than *Escherichia coli* or other Gram negatives [19]. *K. pneumoniae* outbreaks in neonatal care units are well documented in the literature, including the use of pathogen whole-genome sequencing (WGS) to resolve transmission patterns and identify infection sources [20–25]. Nosocomial transmission in LMICs have been linked to poor IPC practices often resulting from limited resources, including inadequate access to water, sanitation, and hygiene, ineffective ward cleaning and sharing of beds and equipment [26–28]. In a retrospective WGS study of *K. pneumoniae* bloodstream infections over 20 years in a single hospital in Malawi, the majority (77%) of cases due to key lineages investigated (ST14, ST15, ST35, ST39) were attributable to nosocomial transmission, suggesting that more effective IPC practices supported by investment in IPC resources could help limit the incidence of neonatal sepsis [22].

To our knowledge, no study has quantified the contribution of nosocomial transmission to the burden of hospital-based *K. pneumoniae* neonatal sepsis across LMICs. This information is crucial to understand and contextualise the role of effective IPC measures in lowering the burden of neonatal sepsis in these settings, which is increasingly important as rising AMR further reduces treatment options. To address this gap, this study aimed to estimate the proportion of *K. pneumoniae* neonatal sepsis infections that are attributable to nosocomial transmission in neonatal care settings in LMICs in Africa and Asia.

## Methods

### Ethical considerations

Each contributing study obtained local ethical approval, details of ethics committees and approval numbers are listed in **S1 Table**. The Baby GERMS-SA [29] and Malawi-Liverpool-Wellcome (MLW) Biobank [22] studies were granted consent waivers from local ethics committees for the use of routine diagnostic specimens/isolates and routine clinical data for research; all other studies obtained informed consent from the parents of participating neonates. Approval for the cross-study analysis presented here was granted by the Observational / Interventions Research Ethics Committee of the London School of Hygiene and Tropical Medicine (ref #29931). Anonymised data from each primary study were shared for analysis, including date of specimen collection and hospital site identifier (where the study included more than one site), along with pathogen WGS data for the corresponding bacterial isolate.

### Bacterial isolates and sequence analysis

Whole-genome sequences of *K. pneumoniae* isolated from neonates in LMICs in Asia and Africa were sourced from prospective clinical studies of neonatal sepsis: Burden of Antibiotic Resistance in Neonates from Developing Societies (BARNARDS) [5], Sepsis Prevention in Neonates in Zambia (SPINZ) [30–32], Baby GERMS-SA [29,33], Mortality from Bacterial Infections Resistant to Antibiotics (MBIRA) [34], Indian District Hospitals (DH) [35], Group B Strep Correlates of Protection Study (GBS-COP) [36], Neonatal Infections & MicroBIome (NIMBI-plus) [37], NeoBAC [38]; and long-term prospective surveillance of bloodstream infection at Queen Elizabeth Central Hospital in Blantyre, Malawi (MLW Biobank) [22] and Kilifi County Teaching and Referral Hospital (KCTRH) carried out by the Kenya Medical Research Institute (KEMRI)/Wellcome Trust Research Programme (KWTRP) [38]. All included studies were undertaken as observational or surveillance studies aiming to capture all blood culture-positive sepsis cases during a defined period, not specifically instigated to investigate or respond to outbreaks. These studies and the corresponding whole-genome sequence data are a subset of those included in a meta-analysis of K and O antigen prevalence amongst *K. pneumoniae* causing neonatal sepsis in LMICs published in 2026 [39]. That study adjusted for within-unit clustering to minimise the impact of localised transmission events on the estimation of regional antigen prevalence, but did not investigate transmission as a phenomenon, which is the goal of the present study. Details of samples included from each study are given in **Table 1**, and characteristics of each study site are given in **Table 2** (this represents information provided by co-authors from each study team). We used the United Nations Statistics Division M49 standard to assign countries to geographical sub-regions (Asian countries) and intermediate regions (African countries) [40].

**Table 1.**
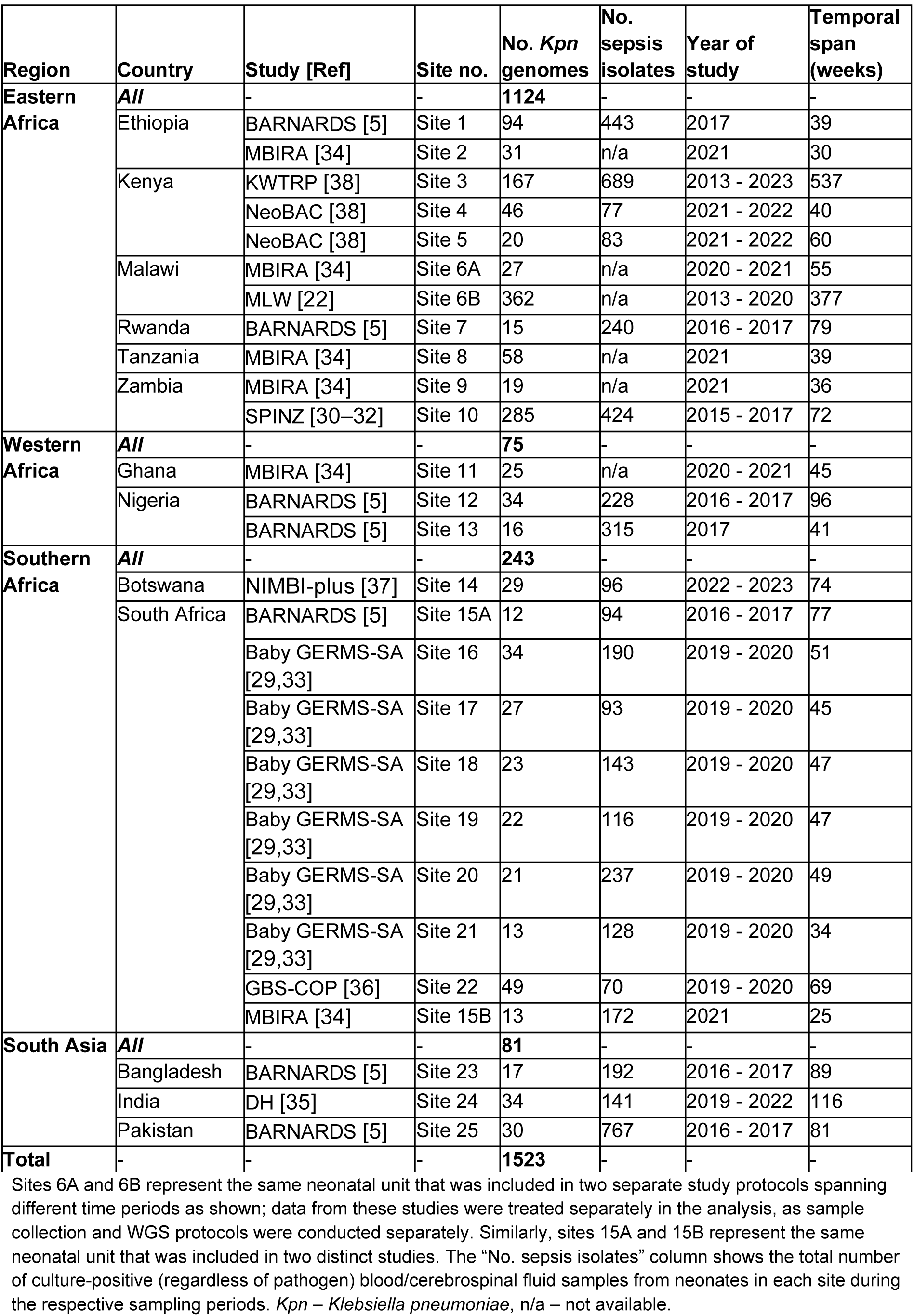
Summary of isolates included in the study.

| Region | Country | Study [Ref] | Site no. | No. *Kpn* genomes | No. sepsis isolates | Year of study | Temporal span (weeks) |
| --- | --- | --- | --- | --- | --- | --- | --- |
| **Eastern Africa** | **All** | - | - | **1124** | - | - | - |
|  | Ethiopia | BARNARDS [5] | Site 1 | 94 | 443 | 2017 | 39 |
|  |  | MBIRA [34] | Site 2 | 31 | n/a | 2021 | 30 |
|  | Kenya | KWTRP [38] | Site 3 | 167 | 689 | 2013 - 2023 | 537 |
|  |  | NeoBAC [38] | Site 4 | 46 | 77 | 2021 - 2022 | 40 |
|  |  | NeoBAC [38] | Site 5 | 20 | 83 | 2021 - 2022 | 60 |
|  | Malawi | MBIRA [34] | Site 6A | 27 | n/a | 2020 - 2021 | 55 |
|  |  | MLW [22] | Site 6B | 362 | n/a | 2013 - 2020 | 377 |
|  | Rwanda | BARNARDS [5] | Site 7 | 15 | 240 | 2016 - 2017 | 79 |
|  | Tanzania | MBIRA [34] | Site 8 | 58 | n/a | 2021 | 39 |
|  | Zambia | MBIRA [34] | Site 9 | 19 | n/a | 2021 | 36 |
|  |  | SPINZ [30–32] | Site 10 | 285 | 424 | 2015 - 2017 | 72 |
| **Western Africa** | **All** | - | - | **75** | - | - | - |
|  | Ghana | MBIRA [34] | Site 11 | 25 | n/a | 2020 - 2021 | 45 |
|  | Nigeria | BARNARDS [5] | Site 12 | 34 | 228 | 2016 - 2017 | 96 |
|  |  | BARNARDS [5] | Site 13 | 16 | 315 | 2017 | 41 |
| **Southern Africa** | **All** | - | - | **243** | - | - | - |
|  | Botswana | NIMBI-plus [37] | Site 14 | 29 | 96 | 2022 - 2023 | 74 |
|  | South Africa | BARNARDS [5] | Site 15A | 12 | 94 | 2016 - 2017 | 77 |
|  |  | Baby GERMS-SA [29,33] | Site 16 | 34 | 190 | 2019 - 2020 | 51 |
|  |  | Baby GERMS-SA [29,33] | Site 17 | 27 | 93 | 2019 - 2020 | 45 |
|  |  | Baby GERMS-SA [29,33] | Site 18 | 23 | 143 | 2019 - 2020 | 47 |
|  |  | Baby GERMS-SA [29,33] | Site 19 | 22 | 116 | 2019 - 2020 | 47 |
|  |  | Baby GERMS-SA [29,33] | Site 20 | 21 | 237 | 2019 - 2020 | 49 |
|  |  | Baby GERMS-SA [29,33] | Site 21 | 13 | 128 | 2019 - 2020 | 34 |
|  |  | GBS-COP [36] | Site 22 | 49 | 70 | 2019 - 2020 | 69 |
|  |  | MBIRA [34] | Site 15B | 13 | 172 | 2021 | 25 |
| **South Asia** | **All** | - | - | **81** | - | - | - |
|  | Bangladesh | BARNARDS [5] | Site 23 | 17 | 192 | 2016 - 2017 | 89 |
|  | India | DH [35] | Site 24 | 34 | 141 | 2019 - 2022 | 116 |
|  | Pakistan | BARNARDS [5] | Site 25 | 30 | 767 | 2016 - 2017 | 81 |
| **Total** | - | - | - | **1523** | - | - | - |

Sites 6A and 6B represent the same neonatal unit that was included in two separate study protocols spanning different time periods as shown; data from these studies were treated separately in the analysis, as sample collection and WGS protocols were conducted separately. Similarly, sites 15A and 15B represent the same neonatal unit that was included in two distinct studies. The “No. sepsis isolates” column shows the total number of culture-positive (regardless of pathogen) blood/cerebrospinal fluid samples from neonates in each site during the respective sampling periods. *Kpn* – *Klebsiella pneumoniae*, n/a – not available.

**Table 2.** Characteristics of facilities included in the study.

| Region | Country | Location | Facility size | No. neonatal beds | Piped water available | Onsite neonatal surgical facilities | Site |
| --- | --- | --- | --- | --- | --- | --- | --- |
| Eastern Africa | Ethiopia | Urban | Very Large | 30 | Sometimes | Yes | Site 1 |
|  |  | Urban | Medium | 40 | Sometimes | Yes | Site 2 |
|  | Kenya | Rural | Large | 75 | Always | Yes | Site 3 |
|  |  | Urban | Large | 100 | Always | No | Site 4 |
|  |  | Urban | Large | 90 | Always | No | Site 5 |
|  | Malawi | Urban | Very Large | 70 | Mostly | Yes | Site 6A/6B |
|  | Rwanda | Rural | Medium | 15 | Always | Yes | Site 7 |
|  | Tanzania | Urban | Very Large | 100 | Always | Yes | Site 8 |
|  | Zambia | Urban | Large | 57 | Mostly | Yes | Site 9 |
|  |  | Urban | Very Large | 60 | Sometimes | Yes | Site 10 |
| Western Africa | Ghana | Urban | Very Large | 55 | Always | Yes | Site 11 |
|  | Nigeria | Urban | Very Large | 30 | Mostly | Yes | Site 12 |
|  |  | Urban | Large | 20 | Always | Yes | Site 13 |
| Southern Africa | Botswana | Urban | Large | 36 | Mostly | Yes | Site 14 |
|  | South Africa | Urban | Very Large | 132 | Always | Yes | Site 15A/15B |
|  |  | Urban | Very Large | 71 | Always | Yes | Site 16 |
|  |  | Urban | Large | 36 | Always | Yes | Site 17 |
|  |  | Urban | Large | 72 | Always | Yes | Site 18 |
|  |  | Urban | Large | 45 | Always | Yes | Site 19 |
|  |  | Urban | Large | 60 | Always | Yes | Site 20 |
|  |  | Urban | Large | 92 | Always | Yes | Site 21 |
|  |  | Urban | Very Large | 200 | Always | Yes | Site 22 |
| South Asia | Bangladesh | Urban | Very Large | 80 | Mostly | Yes | Site 23 |
|  | India | Urban | Small | 12 | Always | No | Site 24 |
|  | Pakistan | Urban | Large | 30 | Always | Yes | Site 25 |

Facility size (total number of beds) – Small: 50–150 beds, Medium: 151–300 beds, Large: 301–600 beds, Very Large: >600 beds

For the prospective studies (BARNARDS, SPINZ, Baby GERMS-SA, MBIRA, DH, GBS-COP, NIMBI-plus, NeoBAC), blood culture was performed for all neonates with clinically suspected sepsis, and all blood culture isolates identified as *Klebsiella* that could be later revived for DNA extraction were included in sequencing and this analysis. Baby GERMS-SA, GBS-COP, KWTRP, and MLW also included *Klebsiella* cultured from cerebrospinal fluid (CSF) of participants. For the long-term prospective surveillance studies (KWTRP, MLW), isolates from all positive blood or CSF cultures were stored for future research. At KWTRP, all neonates had blood culture on admission and again if their clinical condition deteriorated. At MLW, cultures were performed for all neonates with clinical signs of sepsis or meningitis, temperature >37.5°C, or other signs of clinical deterioration.

For all studies, neonatal isolates identified as *Klebsiella*, which could later be revived for DNA extraction, were sequenced and those sequences that passed quality filters were included in this analysis (see inclusion criteria below). Details of microbiology, sequencing, and bioinformatics methods used in each study, including the quality-control criteria applied prior to sharing genome sequences for this analysis, have been reported previously [39] and are summarized in **S2 Table**. Briefly, all studies utilized Illumina platforms and assembled genomes using either SKESA v2.3.0 (MBIRA) or SPAdes (all other studies). Genome assemblies were analysed using Kleborate v3.0 [41] to confirm species, identify multi-locus sequence types (STs), antimicrobial resistance (AMR) and hypervirulence determinants, and to identify capsular (K) loci (KL) and O types using Kaptive v3.0 [42].

### Inclusion and exclusion criteria

Inclusion criteria for individual samples from the studies were: *K. pneumoniae* isolated from the blood or CSF of a neonate (defined as 0-28 days post birth), between 2013 and 2023 inclusive. CSF isolates were included from three studies (KWTRP surveillance, n=5; GBS-COP, n=2; MLW Biobank, n=31). Repeat *K. pneumoniae* isolates from the same participant with the same KL were excluded, ensuring that each genome represents a unique infection from an individual neonate (with the exception of MLW Biobank, for which archived isolates were not associated with patient identifiers thus precluding the exclusion of repeat isolates). Genome sequences identified as species other than *K. pneumoniae* or failing to meet the quality control criteria for assemblies (≤1,000 contigs and genome size 5–6.2 Mbp, as reported by Kleborate v3) were excluded. Additionally, study sites contributing fewer than 10 high-quality *K. pneumoniae* genomes that met the inclusion criteria were excluded (54 genomes from 12 sites), as it is not meaningful to estimate a fraction when the denominator is <10. Specifically, data were excluded from twelve sites that participated in multicenter studies (MBIRA, BARNARDS, DH, GBS) but yielded fewer than 10 high-quality *K. pneumoniae* genomes. Seven of these sites had <5 high-quality *K. pneumoniae* genomes each (no clusters as per the definition below); five sites had n=6-8 isolates each, 3 of which had a cluster. The reasons for low counts vary but are likely associated with differences in patient numbers as well as diagnostic stewardship and sensitivity. A flow diagram detailing sample inclusion is shown in **S1 Figure**, including available information on the number of samples that were not stored, could not be revived for DNA extraction, failed sequencing, or did not meet genome quality control thresholds for each contributing study.

### Identification of transmission clusters

Pairwise single nucleotide variant (SNV) distances were calculated using Pathogenwatch, which enumerates differences in a library of 1,972 core genes (totalling 2, 172, 367 bp) [43]. Transmission clusters were defined within each study site based on both the pairwise genetic distances (≤10 SNVs) and temporal distances (≤4 weeks based on date of specimen collection) between isolates. The igraph v1.4.1 R package [44] was used to identify single-linkage transmission clusters by constructing an undirected graph from an edge list of isolate pairs. Each edge in the undirected graph represented a pairwise connection between isolates that were genetically similar (pairwise distance less than or equal to the specified genetic distance threshold), sampled within a specified temporal window (up to the specified temporal distance threshold), and isolated from the same site. Distinct transmission clusters were then defined as connected components of the resulting graph. This results in single-linkage clusters, where each isolate is within the thresholds of both genetic and temporal distance from at least one other isolate in the cluster, but pairs of isolates within the same cluster may be separated by distances greater than the threshold. Therefore clusters can include isolates cultured over any period, as long as the distances between consecutive isolates in the cluster fall below the temporal threshold.

### Estimation of cluster and transmission proportions

We estimated two key parameters – cluster proportion and transmission proportion – as indices of the contribution of transmission to the burden of *K. pneumoniae* neonatal disease in each neonatal care setting. The cluster proportion estimate was calculated as the number of *K. pneumoniae* isolates in clusters, divided by the total number of culture-confirmed and sequenced *K. pneumoniae*, for each site. The transmission proportion, on the other hand, was a conservative estimate of the proportion of *K. pneumoniae* isolates that were attributable to transmission. To estimate this, we excluded the first neonate in each cluster as a potential index patient, and assumed that all other infections within clusters were due to onward transmission in the unit. This almost certainly results in an underestimation of infections attributable to transmission, as the index patient may have also acquired *K. pneumoniae* from a contaminated source within the unit or an asymptomatically colonised neonate. The transmission proportion estimate was thus calculated as below:

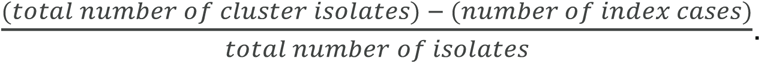

### Sensitivity analyses

We conducted a sensitivity analysis to determine the impact of the choice of genetic distance and temporal distance thresholds on the estimates of cluster proportion and transmission proportion. The estimates were calculated and compared across a range of genetic distance thresholds (range: 0–25 SNVs) and temporal distance thresholds (range: 1–52 weeks). We also assessed how using different methods for calculating pairwise genetic distances between all isolate pairs could impact the obtained estimates. To do this, we generated reference-free SNV alignments of all genomes in the BARNARDS and SPINZ datasets using the split k-mer analysis (SKA) method with SKA2 v0.4.1 [45,46]. A SKA index was first built from the input assemblies using a k-mer size of 31, after which pairwise distances between all genomes were calculated using the “distance” subcommand with a minimum frequency threshold of 0.9 to include only core-genome SNVs. The cluster and transmission proportion estimates were re-calculated using these SKA-derived SNV distances and compared to those obtained using the Pathogenwatch core gene SNV distances.

### Statistical analysis

All analyses and visualisations were performed using R v4.2.3 [47].

The methods for clustering infections, quantifying transmission, and exploring the sensitivity of the estimates to selected thresholds (as described above) are implemented in a species-agnostic Shiny web application (https://klebsiella.shinyapps.io/transmission_estimator/), with source code available at https://github.com/klebgenomics/transmission_estimator (DOI: 10.5281/zenodo.17593948).

To assess heterogeneity of estimates between sites, for the multicentre studies BARNARDS and MBIRA, we performed meta-analysis of proportions using a generalised linear mixed model (GLMM) with a logit transformation to pool estimates of cluster proportion and transmission proportion across all study sites. A random effects model was applied to account for between-site variability, and 95% confidence intervals were computed using the Clopper-Pearson method. Heterogeneity between sites was assessed visually using Forest plots and quantitatively using the *I^2^* statistic. The meta-analysis was conducted using the metaprop function of the meta v7.0.0 R package [48].

To identify facility characteristics associated with transmission, we performed pairwise two-tailed binomial tests with Bonferroni correction, with facility-level data as categorical (facility size, number of neonatal beds, availability of piped water) or binary (onsite availability of neonatal surgical facilities) variables.

The proportions of isolates with extended-spectrum beta-lactamase (ESBL) and carbapenemase genes were compared between clustered and non-clustered cases using the Chi-squared test. To model different bacterial features (ESBL gene carriage, carbapenemase gene carriage, ST, O type, K locus) and institutional factors (hospital site) as potential predictors of transmission, we fitted logistic regression models in which each introduction was treated as a single observation, with the outcome recorded as transmission (cluster size ≥2) or no evidence of transmission (unclustered singleton case). For each cluster, each predictor was assigned the consensus value for that cluster (e.g. if 3 of 4 cases had ESBL gene/s detected, the cluster was recorded as ESBL-positive). Presence of ESBL (Kleborate score ≥1) or carbapenemase (Kleborate score ≥2) genes were encoded as binary variables. ST was encoded as a categorical variable, with one category per ST for commonly transmitted STs (occurring in ≥3 clusters and ≥2 sites), and the reference category being the group of all 158 ‘other STs’ (49.2% in the reference category). O type was encoded as a categorical variable, with the most common type, O1 (51.6%), as the reference category. K locus was encoded as a categorical variable, with the top 10 K loci compared against ‘other K loci’ as the reference category (n=70, 87.5% in the reference category). Models were fitted using Firth’s bias reduction method, implemented in the logistf package in R (v1.26.0). All alternative models were compared using the likelihood ratio test, using the anova function in base R.

## Data availability

Sample-level data are described in **S3 Table** (dates are shown as year only, to protect participant privacy). Raw whole-genome sequence data were deposited by the originating study teams in INSDC databases, under the following BioProjects: BARNARDS, PRJEB33565; SPINZ, PRJEB46513; MLW, PRJEB42462; NIMBI-plus, PRJNA1168993; DH, PRJEB70311; Baby GERMS-SA, PRJNA796486 and PRJNA1282934; GBS-COP, PRJNA1175467; KWTRP, PRJNA1265413; MBIRA: PRJNA1274034, NeoBAC, PRJNA1265413. All input data and code required to reproduce the results, figures, and tables presented in this paper are available at https://github.com/klebgenomics/KlebNNS_transmission (DOI: 10.5281/zenodo.17591910). This study is reported as per the Strengthening the Reporting of Observational Studies in Epidemiology (STROBE) guideline (**S1 Checklist**).

## Results

### Estimating transmission proportion

The study included 1,523 isolates from 27 sites in 13 countries, including Malawi (n=389), Zambia (n=304), Kenya (n=233), Ethiopia (n=125), Tanzania (n=58) and Rwanda (n=15) in Eastern Africa; Nigeria (n=50) and Ghana (n=25) in Western Africa; South Africa (n=214) and Botswana (n=29) in Southern Africa; India (n=34), Pakistan (n=30) and Bangladesh (n=17) in South Asia (**Table 1, S2 Figure**). Sample size per site ranged from 12 to 362 genomes, with dates of isolation spanning 26 to 537 weeks per site. Six sites included data from only one year. A majority of the isolates were from Eastern Africa (n=1124), followed by Southern Africa (n=243), South Asia (n=81), and Western Africa (n=75). See **Table 2** for characteristics of neonatal care facilities included in the study.

A total of 156 transmission clusters were identified across all sites, comprising 1,035 unique infections (68.0%) (**S3 Figure**). The identified clusters spanned between 1 and 249 days duration between the first and last cases (median: 14 days; interquartile range [IQR]: 6–28 days), with a median of 3 infections per cluster (range: 2–188, IQR: 2–5) (**S4 Table)**. Almost half (73/156) of the clusters comprised only 2 infections. Excluding these 73 clusters as potential coincidental isolation of similar but independently introduced strains rather than nosocomial transmission, there were still 879 (57.7%) infections involved in 83 clusters affecting at least 3 patients each (median cluster size, 5 cases). Sixteen clusters overlapped either the beginning (n=7, cluster size: median = 3, range = 2–188) or the end (n=9, cluster size: median = 3, range = 2–32) of the sampling periods in the respective sites, suggesting that transmission may have been ongoing before sampling began or continued after sampling ended.

The median estimates, across 27 sites, for cluster and transmission proportion were 53.3% and 33.5%, respectively (**Figure 1**). Sensitivity analyses showed these estimates were robust to the choice of thresholds and genetic distance estimation methods (**S4 Figure, S1 Appendix**). Cluster and transmission proportion estimates varied widely by site, ranging from 0 to 93% and 0 to 87%, respectively (**Figure 1**). In 10 of the 27 sites (37%), at least half of all infections were attributable to transmission, and in 14 sites (52%) at least a third of cases were attributable to transmission. Across all sites, the median number of clusters detected per site per year was three (IQR: 2–4, range: 0–13). Considering only clusters with ≥3 patients, we identified median one cluster per site per year (IQR: 1–3, range: 0–8). Transmission proportions were heterogeneous within regions, although higher values were estimated in Eastern Africa (median=65.0% [range: 29.6–93.5%], n=11 sites) compared with Western Africa (40% [37.5–41.2%], n=3 sites) and South Asia (43.3% [23.5–88.2%], n=3 sites). Intermediate values were estimated in Southern Africa (median = 54.8% [0–77.3%], n=10 sites).

**Figure 1.**
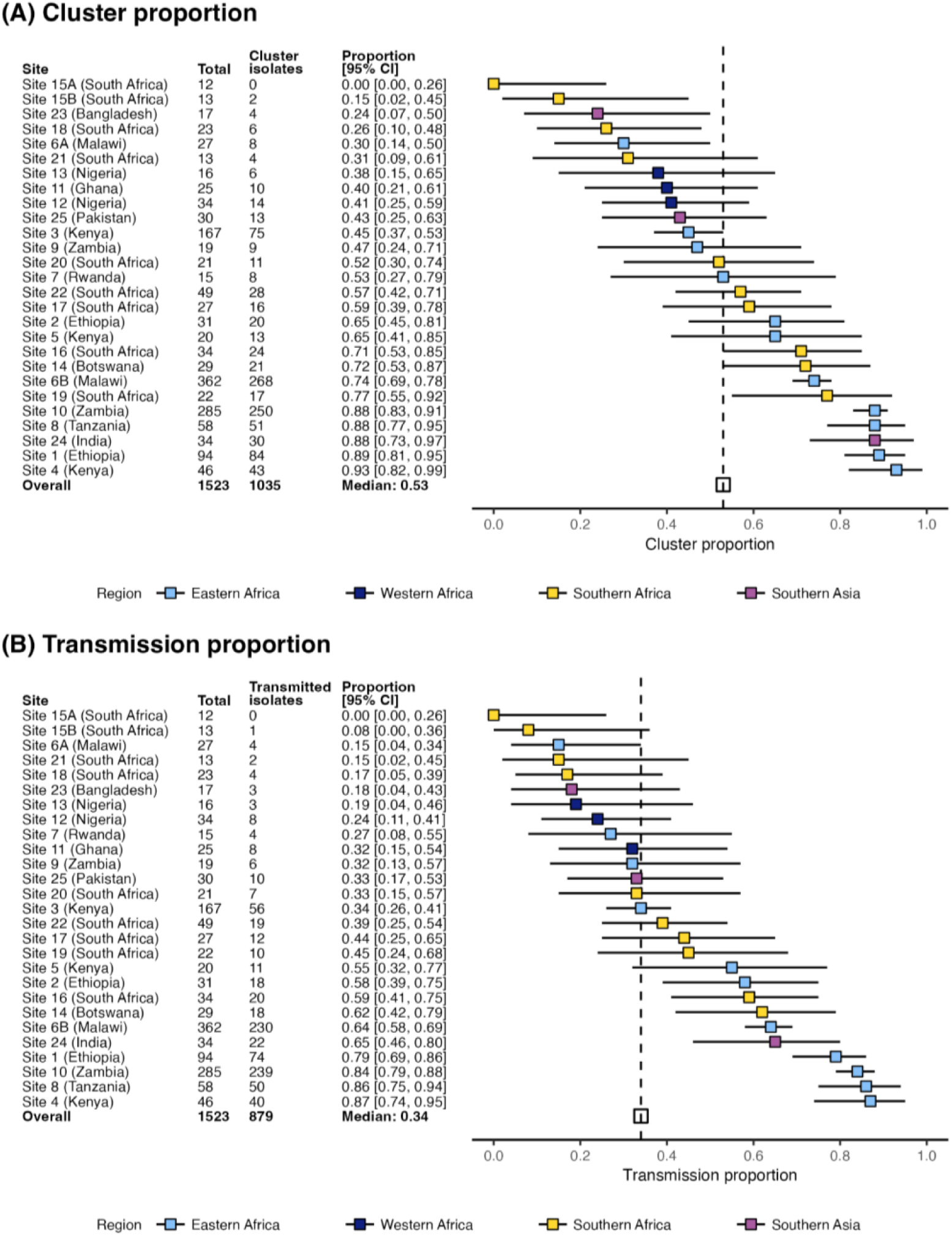
Estimates of cluster and transmission proportion across sites. **(A)** Cluster proportion **(B)** Transmission proportion. Plots show the point estimates (boxes) and 95% confidence intervals (horizontal bars) for each site. The median estimate is represented by the broken vertical line.

To determine whether differences in study design and sampling methodology contributed to the heterogeneity observed between sites, we conducted two secondary meta-analyses using data from two multi-site studies in our dataset (MBIRA and BARNARDS), each of which used a consistent study protocol across multiple sites [5,34]. The pooled estimate of transmission proportion across the seven BARNARDS sites was 26.1% (95% CI: 11.5–49.0%, range: 0–79%) and there was substantial heterogeneity between sites (*I^2^*= 88.6% [79.0%–93.8%]). The findings with the MBIRA dataset were similar, with a pooled transmission proportion estimate of 37.0% (95% CI: 15.8–64.8%, range: 8–86%) and significant heterogeneity (*I^2^* = 89.0% [78.8–94.3%] was observed across the six sites. Notably, the lowest cluster proportion observed in both these studies was at the same neonatal unit, sampled under different protocols at different time periods (BARNARDS 15A, 2016-2017 and MBIRA 15B, 2021), but yielding similar results each time (0 clusters out of 12 isolates, and 1 clustered pair out of 13 isolates, respectively). This suggests that the observed heterogeneity in the full analysis is unlikely to be influenced by primary study protocol differences alone, but may instead reflect variation in unmeasured factors such as demographic, geographic and/or site-specific variations.

**S5 Figure** shows a comparison of cluster proportion across different sites based on descriptive characteristics of the health facilities (summarised in **Table 2**). Notably facility size and neonatal bed count were not associated with the number of *K. pneumoniae* isolates per site (R^2^ <0.01, p=0.993 and R^2^ = 0.14, p=0.053, respectively using linear regression). For example, the lowest cluster proportions were observed at site 15, which is one of the largest neonatal units but as noted above had very few cases or clusters, potentially because it is one of the better-resourced facilities (located in Cape Town, South Africa with relatively strong microbiological diagnostics, surveillance and IPC capacity). Facilities with only occasional availability of piped water (two in Ethiopia, one in Zambia) had a higher proportion of cases in clusters (87.7% [range: 64.5–89.4%]) than those where piped water was reported as being available most of the time (n=6; 42.5% [23.5–74.0%]; p<0.001 using Binomial test) or always (n=18; 52.9% [0–93.5%]; p<0.001 using Binomial test) (**S5 Figure**). District hospitals with no surgical facilities onsite (two in Kenya, one in India) also had a higher proportion of cases in clusters (88.2% [65.0–93.5%]) than hospitals with surgical facilities available onsite (n=24, 49.9% [0–89.4%], p<0.001 using Binomial test).

### Genomic characteristics of transmitted *K. pneumoniae*

A total of 172 distinct *K. pneumoniae* STs were identified amongst the included isolates (**S5 Table**). Of these, 57 STs were associated with at least one transmission cluster. We defined commonly transmitted STs as those that were identified in ≥3 transmission clusters in two or more sites. Fourteen STs met this definition (**Figure 2**), and these made up 63.8% (n=972/1523) of all infections, including 71.8% (n=743/1035) of clustered infections and 46.9% (n=229/488) of unclustered infections. Twelve of the 14 commonly transmitted STs were detected in ≥3 countries each, however ST152 and ST25 were each identified in just one country (South Africa and Malawi, respectively). Most of the 14 STs were associated with clusters at Malawi site 6B (see **Figure 2B**), which is not unexpected given this is the largest dataset and originates from seven years of routine blood culture surveillance; however, besides ST25, all common STs associated with clusters at this site were also associated with clusters at multiple other sites. The 14 commonly transmitted STs were responsible for most large clusters with ≥15 cases (n=6/10 clusters; 60%; **Figure 2D**), although four large clusters were caused by rarely transmitting STs (ST6775, n=29; ST37, n=26; ST2004, n=20; and ST1414, n=17). **Figure 3** shows, for each ST, the number of unique introductions (treating each cluster or singleton as a unique strain ‘introduction’ in a neonatal ward) vs the number of infection clusters. Across all STs, the mean fraction of introductions resulting in detected infection clusters was 15.3%, and this fraction was significantly higher among the 14 commonly transmitted STs compared to the other 158 STs (mean 33.1% vs 13.8%, Chi-squared test p<0.001).

**Figure 2.**
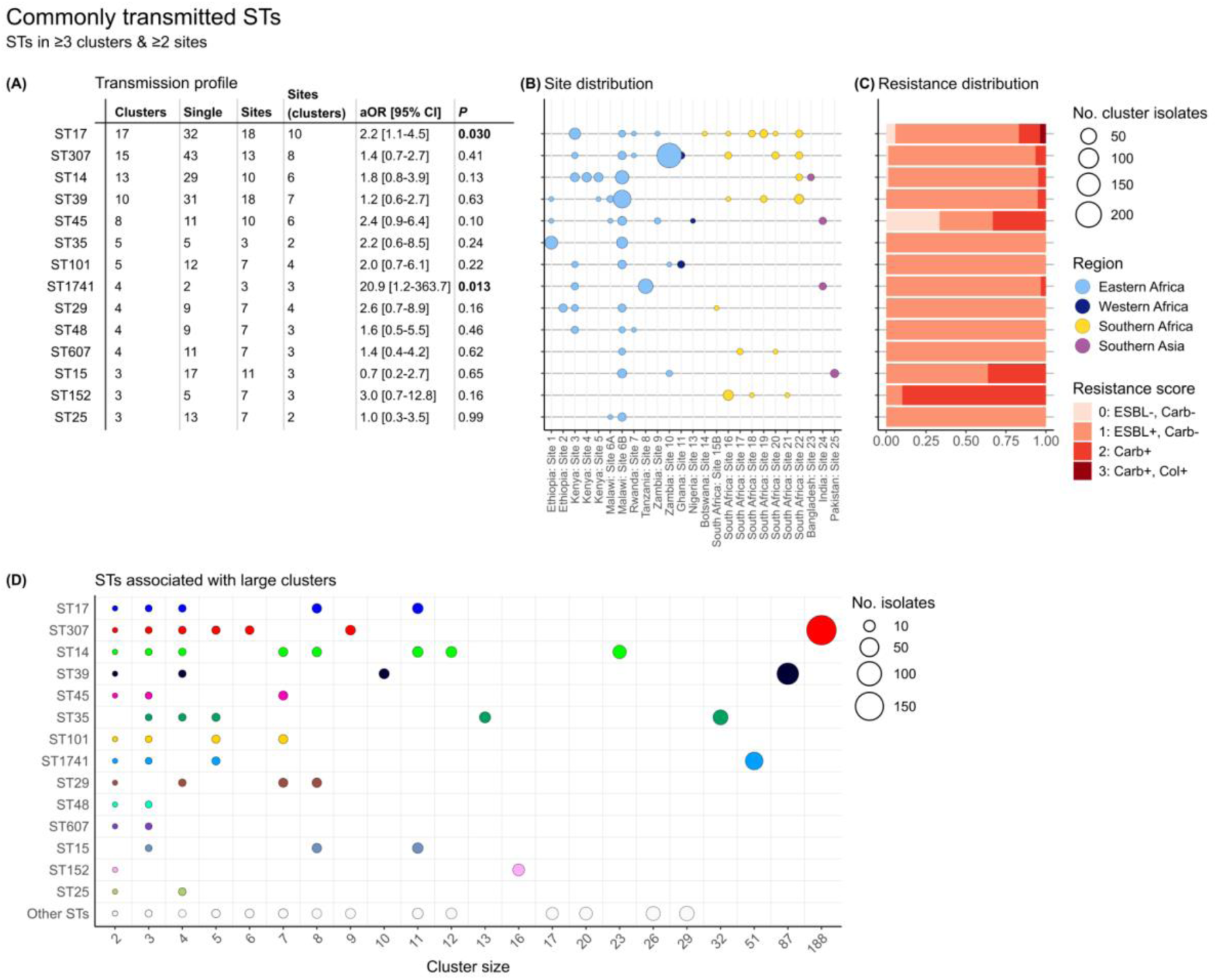
Commonly transmitted *K. pneumoniae* STs. **(A)** Transmission profile showing number of clusters, number of singleton cases, total number of sites the STs were detected, number of sites where the STs were part of transmission clusters, their adjusted odds ratios compared to other STs in a logistic regression model to assess association with transmission, and corresponding p-values (columns from left to right). P-values for STs with a significant association with transmission are formatted in bold (see Methods for details of the logistic regression analysis). **(B)** Site distribution of cluster isolates. Points are coloured to indicate geographical region and sized to indicate number of clustered isolates per site. **(C)** Resistance distribution (proportion) of cluster isolates. Resistance scores are as reported by Kleborate based on the presence or absence of extended-spectrum beta-lactamase genes (ESBL), carbapenemase genes (Carb), and/or colistin resistance determinants (Col). **(D)** ST composition of large clusters. Points are coloured to indicate the 14 commonly transmitted STs and sized to indicate the number of isolates, as per the figure legend.

**Figure 3.**
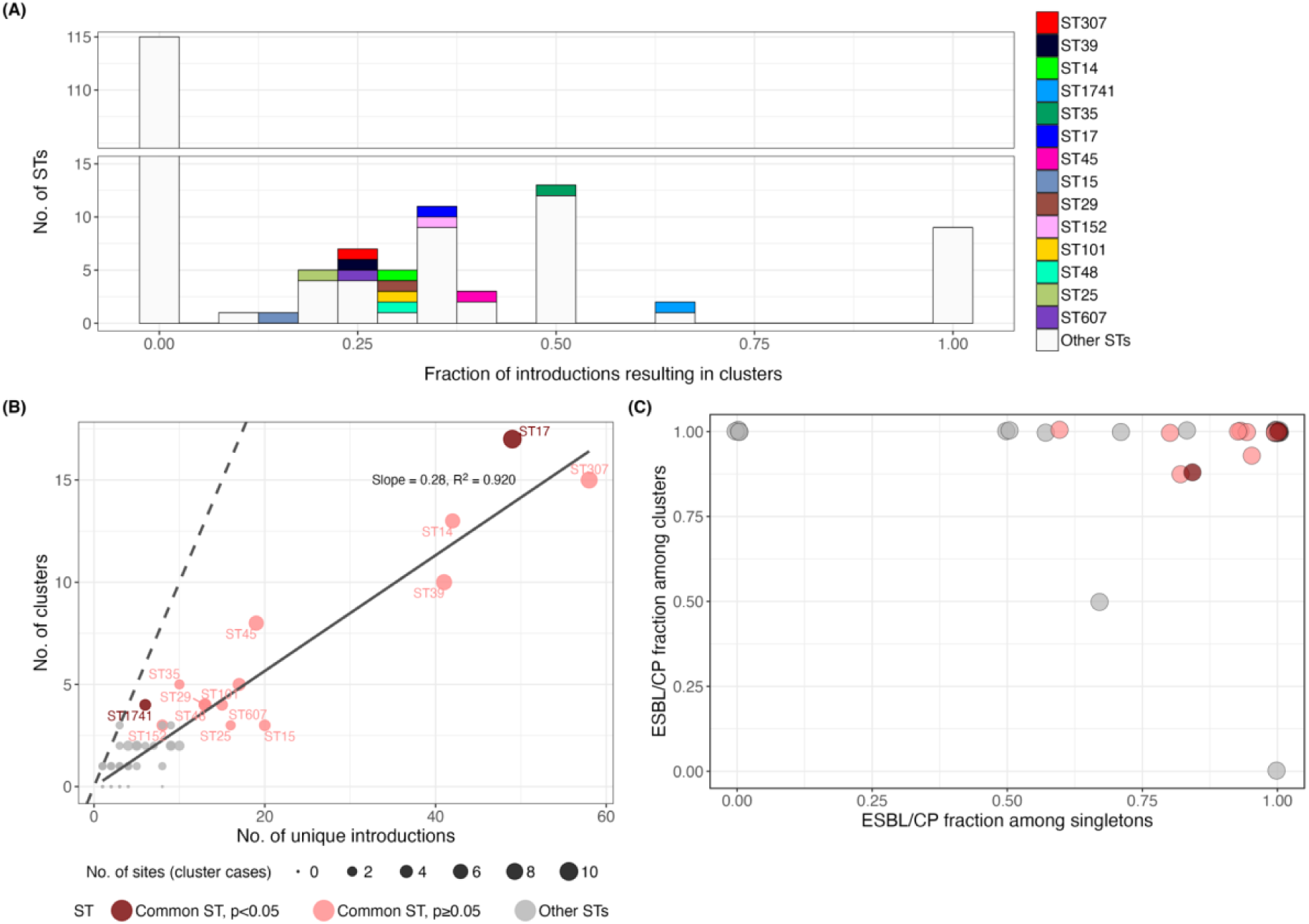
Transmission probability of individual STs. **(A)** Distribution of the fraction of strain introductions resulting in clusters, for individual STs. Bars are coloured to indicate commonly transmitted STs (per the panel legend), defined as STs that were identified in ≥3 transmission clusters in two or more sites. **(B)** Number of unique introductions vs number of clusters, for individual STs. Points are sized to indicate the number of sites where clustered isolates were detected for each ST, and coloured to indicate commonly transmitted STs significantly associated with transmission in the logistic regression model. The dashed line shows y=x, along which the number of unique introductions is equal to the number of clusters. The solid line represents a linear regression line of best fit through the origin (slope 0.28, R^2^=0.920). **(C)** Fraction of clusters and singletons with ESBL and/or carbapenemase. Points are plotted for the 57 STs involved in clusters, and coloured to indicate commonly transmitted STs significantly associated with transmission. ST: sequence type. ESBL/CP: extended-spectrum beta-lacatamase or carbapenemase gene carriage (Kleborate resistance score ≥ 1).

The majority of isolates (n=1384/1523, 90.9%) carried an ESBL or carbapenemase gene. Clusters were more likely to carry ESBL (94.2% of clusters (n=147/156) vs 81.8% of singletons (n=399/488), p<0.001) or carbapenemase genes (19.9% of clusters (n=31/156) vs 10.2% of singletons (n=50/488), p=0.0025) (**S6 Table**). In a logistic regression model for transmission, treating each cluster or singleton as a unique strain ‘introduction’ in a neonatal ward, and defining the outcome for each introduction as either evidence of transmission (i.e. cluster of size ≥2) or no evidence of transmission (i.e. a singleton unclustered case), transmission was significantly positively associated with carriage of ESBL genes (adjusted odds ratio, aOR = 2.84 [95% CI: 1.45–5.57], p<0.001) or carbapenemase genes (aOR = 1.81 [95% CI: 1.10–2.98], p=0.02) (**S7 Table**). Including Site as a covariate improved model fit (p=0.05 using likelihood ratio test), but did not substantially alter the effect estimates for ESBLs (aOR = 2.48 [95% CI: 1.26–4.90], p=0.006) or carbapenemases (aOR = 2.08 [95% CI: 1.04–4.14], p=0.04).

It is challenging to tease apart the effect of AMR from the effect of lineage, as certain lineages may appear more likely to cause detectable infection clusters due to their resistance genes (if AMR is a driver of transmission and/or infection), or strains with resistance genes might appear more likely to cause detectable infection clusters due to their association with particular genetic lineages (if non-AMR-related lineage-associated differences drive transmission and/or infection). The 14 commonly transmitted STs showed a significantly higher rate of ESBL gene carriage (amongst singleton infections or deduplicated clusters) compared to the other 158 STs (94.2% (n=308/327) vs 75.1% (n=238/317), p<0.0001), although there was no significant difference in carbapenemase gene carriage rates (14.1% (n=46/327) vs 11% (n=35/317), p=0.30) (**S8 Table**). Amongst the 14 most common transmission STs, nearly all (n=728/743; 98%) of the clustered isolates carried at least one ESBL or carbapenemase gene, compared with 93% (n=213/229) amongst singleton isolates of these STs (**Figure 3C**). Adding ST (as a categorical variable with each of the 14 common STs as unique categories, compared to all other STs as the reference category) to the logistic regression model with ESBL, carbapenemase and Site did not improve the model (p=0.226), and had little impact on the estimated effects of ESBL (aOR = 2.16 [95% CI: 1.08–4.32], p=0.03) and carbapenemase (aOR = 2.03 [95% CI: 1.01–4.08], p=0.05), and only two STs were significantly independently associated with transmission (ST17, aOR = 2.24 [95% CI: 1.11–4.52], p=0.03 and ST1741, aOR = 20.9 [95% CI: 1.2–363.7], p=0.01).

As K and O antigens have been proposed as potential vaccine targets, we explored their prevalence (based on genomic predictions) and association with transmission in our dataset. Six unique O serogroups were predicted (see **S9 Table**), the majority being O1 (n=332/644, 51.6%) or O2 (n=158/644, 24.5%), which were common amongst both clusters and singletons (51.3% vs 51.6%, respectively for O1, 26.9% vs 23.8% for O2). Including O types in the transmission model with ESBL and carbapenemase genes, with or without ST and/or Site, did not improve model fit (p>0.2 using likelihood ratio test), and no O types were significant in any models (see **S7 Table**).

Eighty distinct K loci were detected. Forty-three K loci were found in isolates associated with transmission clusters, and 39 of these were also identified in singleton isolates (**S10 Table**). Distinct K loci were associated with specific STs, making it difficult to tease apart the effect of K antigens from strain background. For example, KL102, which was the most common K locus associated with transmission, was closely associated with ST307: 95% (n=241/253) of KL102 isolates from transmission clusters were ST307, and ST307 accounted for 83% (n=15/18) of KL102 clusters detected. Similarly, 79% (n=42/53) of KL25 cluster isolates belonged to either ST17 (n=33, 62%) or ST607 (n=9, 17%). Including the top 10 most common K loci in the transmission model together with ESBL and carbapenemase genes, with or without ST and/or Site, did not improve the model fit (p>0.38 using likelihood ratio test), although KL62 was significantly positively associated with transmission (aOR 4.77 [95% CI, 1.30–17.49], p=0.025) in the model with ESBL and carbapenemase genes, ST, and Site. KL62 was associated with 12 clusters (n=26 isolates) out of 34 introductions (n=6 different STs) across 16 sites.

## Discussion

Neonatal sepsis caused by *K. pneumoniae* infection is a significant health challenge in Africa and Asia with disproportionately high incidence and mortality rates. Our analysis estimated that over half of all neonatal sepsis cases at the included sites were within transmission clusters, with a median 33.5% of all cases attributable to nosocomial transmission (**Figure 1**).

Transmission analyses typically require detailed data on patient admission, ward movements, and recent healthcare exposures in addition to pathogen sequence data; these are hard to standardise across studies and thus between-study comparisons or meta-analyses are challenging. Here, however, we used studies in which all consecutive neonates with sepsis in a defined physical unit were sampled and the resulting isolates sequenced, allowing us to undertake a standardised clustering analysis based on genetic and temporal distances within each unit, applied to data collected across 10 unique studies.

Furthermore, as our goal was to estimate the fraction of cases linked to others in transmission clusters, rather than trying to resolve specific transmission chains or pathways, our approach was robust to changes in distance thresholds (see **S4 Figure, S1 Appendix**). This is because lowering clustering thresholds tends to break larger single-linkage clusters into smaller ones (which has no impact on the overall fraction included in clusters), rather than breaking clusters into completely un-linked singleton cases. Variations in genetic and temporal distance thresholds have a greater impact in real-time outbreak investigations, where the goal is to identify specific transmission events and sources. In that context, the choice of thresholds can be adjusted to achieve a desired preference for sensitivity or specificity in cluster detection. Interestingly, previous studies have proposed whole-genome SNV cutoffs of between 21 and 25 for identifying *K. pneumoniae* nosocomial transmission, which is equivalent to the 10 Pathogenwatch core-gene SNV threshold used in our analysis [18,49,50].

As with other opportunistic pathogens frequently implicated in hospital-acquired infections, reservoirs and transmission routes for *K. pneumoniae* infection are abundant in the hospital environment, and genomics has been used to resolve specific sources in several studies. These include transmission through direct contact with contaminated surfaces, sinks, cleaning supplies, medical devices, and healthcare personnel [51–55]. Neonates are at high risk of hospital-acquired infections due to their naive or compromised immune systems, exposure through invasive procedures in critical care settings, repeated handling by multiple caregivers, and long hospital admissions [56,57]. While our study indicates a high impact of IPC deficiencies in neonatal units in LMICs, identifying suitable solutions will require better understanding of risk factors, transmission routes, and sources of contamination. The solutions will also require appropriate testing in clinical trials or complex interventions and subsequently funding for implementation at scale.

We observed a high diversity of *K. pneumoniae* lineages (172 unique STs) implicated in neonatal sepsis cases across hospitals in Asia and Africa (**S5 Table**), consistent with previous reports of a highly diverse etiology of *K. pneumoniae*-associated neonatal sepsis [5,22]. A third of these STs were part of at least one transmission cluster, among which 14 STs were identified as commonly transmitted, occurring in ≥3 clusters and ≥2 sites. However, most of these were also common among singleton isolates and accordingly our logistic regression analysis indicated only two of these STs were significantly associated with transmission clusters, compared with other STs. This finding is consistent with the hypothesis that many different STs can become common causes of transmission clusters, either because they possess traits that render them particularly fit for transmission in the hospital setting, or simply because they are more prevalent (e.g. as gut colonisers in the community) and therefore have greater stochastic opportunity due to more frequent introductions to the hospital setting. Importantly, the signal of transmission we are exploring in this study – detection as a cluster of clinical infections – is the product of multiple factors including colonisation potential, virulence, and environmental survival, and we are unable to tease apart the relative contributions of these different factors in determining which strains are identified frequently in clusters. Notably, many of the 14 commonly transmitted STs identified here, including ST11, ST14, ST15, ST17, ST29, ST101 and ST307 are recognised high-risk clones, widely implicated in multidrug-resistant (MDR) hospital-acquired infections worldwide [58–60], consistent with our observations. ST39 has been reported less frequently, but is also disseminating rapidly worldwide, including in Africa, where it has been implicated in MDR neonatal outbreaks in The Gambia and Malawi [22,28,61].

The overall high rates of AMR among the *K. pneumoniae* isolates (clustered and singleton) in this study (see **S6 Table**) is concerning, as a high degree of resistance is known to be associated with higher rates of nosocomial spread [18,62], a finding that was corroborated in our analysis. We found that cluster cases had significantly higher rates of ESBL and carbapenemase gene carriage compared to single cases, and isolates belonging to the 14 most commonly transmitted STs more frequently carried ESBL genes compared to other STs (**S6 Table, S8 Table**). Carbapenemase and ESBL gene carriage were also independently associated with transmission in the multivariable regression analysis (**S7 Table**). It is likely that these AMR phenotypes also contribute to case detection, as strains resistant to empirical therapy are more likely to be present in the blood at sufficient quantities to be detected by blood culture, making it more likely for transmitted AMR strains to result in cluster detection than transmitted susceptible strains.

While AMR almost certainly plays a role in the successful persistence of *K. pneumoniae* clones within the hospital setting, other potential factors such as survival on environmental surfaces and colonisation ability are less well characterised [63]. For instance, ST17, the most commonly transmitted ST in this study, was recently demonstrated to colonise the gastrointestinal tracts of children for up to 2 years, a factor that may contribute to its increased transmissibility [64]. ST307 is frequently associated with outbreaks and believed to be adapted to the hospital environment where it can persist for extended periods and rapidly spread among patients [25,63,65]. In this study, ST307 was responsible for the largest identified cluster (involving 188 neonates over a 32-week period at a hospital in Zambia), and 14 other clusters across 8 sites in Kenya, Malawi, Rwanda, Zambia, South Africa and Ghana. However, ST307 was also detected in 43 singleton infections, and only 26% (n=15/58) of detected ST307 introductions in the neonatal units were associated with transmission clusters (similar to the background rate observed across 158 non-commonly transmitted STs). This suggests that the frequency of ST307 among hospital outbreaks may reflect a generally high prevalence (i.e., as a gut coloniser) resulting in greater opportunity, in combination with or instead of a specific propensity for transmission in the hospital setting. Further research is needed to determine if specific STs have enhanced risk for transmission once introduced into the hospital setting, and to elucidate potential factors that contribute to their success as hospital-acquired opportunistic pathogens.

In summary, our findings demonstrate the need to improve IPC practices in neonatal care settings in LMICs, and highlight the potential benefits of genomic surveillance of *K. pneumoniae* and other pathogens to assess IPC impacts and/or to support real-time outbreak detection and investigation. Limiting horizontal transmission through improved IPC measures is undoubtedly a crucial prevention strategy that could prevent a significant proportion of neonatal sepsis caused by *K. pneumoniae* and other pathogens, and curb the spread of AMR. Studies have shown that *K. pneumoniae* outbreaks in neonatal units can in principle be effectively controlled by implementing simple IPC measures like contact precautions and improved hand hygiene [21,28,31]. However, to be truly effective in reducing overall disease burden, these measures need to be proactive, rather than reactive, and be integrated into routine practice. The implementation of consistent and effective IPC measures in resource limited settings remains challenging due to resource constraints, inadequate infrastructure, limited cleaning supplies, understaffing, and limited access to training [66,67]. Whilst this analysis was not suited to probing the determinants of variation in transmission rates, we did find that the three facilities with limited access to piped water displayed high cluster proportions, highlighting the challenges faced in some settings and their impact on hygiene and patient safety. In contrast, the lowest cluster proportions and the lowest isolate counts were observed in consecutive studies at the same large tertiary care hospital in South Africa, which is comparatively well resourced in terms of infrastructure and staffing. Simplified IPC models tailored to resource-constrained settings and focused on training, improved cleaning, and hand hygiene have been proposed and demonstrated to reduce the incidence of neonatal sepsis and mortality [31]. However it is clear that to be effective, such measures must be supported by sustained investment in infrastructure, equipment, staff and training [68]. In addition, to maximise their effectiveness, IPC implementation should be periodically assessed using suitable assessment tools and reviewed as necessary [69–71]. IPC strategies must also be supported by expanding access to blood culture facilities [72,73], accurate diagnostics and robust clinical and microbiological surveillance [74], ideally incorporating whole-genome sequencing, to enable prompt detection of infections, precise transmission tracking, and timely institution of control measures.

Additionally, our findings also have implications for clinical management. WHO guidelines for empirical treatment of neonatal sepsis (third-generation cephalosporins plus gentamicin [75]) are likely to be ineffective against the vast majority of *K. pneumoniae* characterised in this study, which mostly carried ESBL genes, consistent with recent reports from NeoOBS and other studies [11,76]. Access to blood culture facilities, robust clinical diagnostics and susceptibility testing is essential to monitor AMR and update local syndromic guidelines, as is improving access to effective antibiotics in all settings.

### Limitations

The major limitation of this study is the unavailability of standardised clinical and patient-level data across the included sites, which precludes a more in-depth investigation of potential transmission routes and overall transmission dynamics, and the delineation of infections based on the timing of disease onset relative to admission (which can be an indicator of hospital- vs community-acquired infection) or to date of birth (early vs late onset sepsis). Notably, component studies with sufficient data to investigate onset relative to admission showed that clustered cases often had rapid onset following admission to the neonatal unit, and one study linked rapid-onset disease to contaminated intravenous fluids, confirming nosocomial transmission [32,37]. This suggests that the timing of symptom onset following admission is not a reliable indicator of hospital- vs community-acquired disease in these settings, even if such data were available. This study also did not include matched maternal data to investigate vertical transmission as an alternative transmission route, albeit it is important to note that studies suggest that this is unlikely to play a significant role in *K. pneumoniae* neonatal sepsis [5,77]. Another limitation was the reliance on *post hoc* collaborator-elicited facility-level data rather than systematically collected data to explore the potential causes of variation in transmission rates across the different hospitals. Further studies using tailored, prospectively collected data are needed to explore site-specific factors that may influence transmission risk. Also, as this study predominantly comprised tertiary-level healthcare facilities, our findings may not be generalisable to lower-tier healthcare facilities.

The attributable transmission fraction observed in our analysis (median = 33.5%) is likely an underestimate due to the conservative nature of our transmission proportion definition. Our assumption that the ‘index cases’ were not attributable to transmission discounts the very likely possibility that they themselves acquired their infection from the same source as the subsequent cases (e.g., via contaminated equipment or the hands of a carer). The cluster proportion is also likely biased towards underestimation due to missed cases (including colonisation episodes that did not result in a culture-confirmed infection), excluded cases (e.g., children aged >28 days in the same neonatal units), or cases that were acquired in the neonatal unit but not clustered with another sequenced isolate (e.g. a contaminated source may lead to colonisation or even disease in multiple neonates, of which only one individual case of invasive disease is detected). Given these underestimations, even the cluster proportion (median = 53.3%) probably provides a conservative estimate of the true fraction of sepsis cases amenable to prevention with improved IPC measures.

## Supporting information

S1 Checklist

## Data Availability

Sample-level data are described in Table S3 (dates are shown as year only, to protect participant privacy). Raw whole-genome sequence data were deposited by the originating study teams in INSDC databases, under the following BioProjects: BARNARDS, PRJEB33565; SPINZ, PRJEB46513; MLW, PRJEB42462; NIMBI-plus, PRJNA1168993; DH, PRJEB70311; Baby GERMS-SA, PRJNA796486 and PRJNA1282934; GBS-COP, PRJNA1175467; KWTRP, PRJNA1265413; MBIRA: PRJNA1274034, NeoBAC, PRJNA1265413. All input data and code required to reproduce the results, figures, and tables presented in this paper are available at https://github.com/klebgenomics/KlebNNS_transmission (DOI: 10.5281/zenodo.17591910)

## Acknowledgments

We thank the participants and their families in all the contributing studies, and the clinical and laboratory staff involved in collection and processing of relevant samples and isolates. This work was supported by the MASSIVE HPC facility (www.massive.org.au). We would also like to acknowledge the expert support of the sequencing and computational genomics teams at the CHOP Microbiome Core (Philadelphia), International Livestock Research Institute (Nairobi), Kilimanjaro Clinical Research Institute (Moshi), Neuberg Center for Genomic Medicine (Ahmedabad), NICD (Johannesburg), Quadram Institute (Norwich), Wellcome Centre for Human Genetics (Oxford), and Wellcome Sanger Institute (Cambridge); and the BARNARDS microbiology and genomics team at Cardiff University.

## Supporting Information Captions

**S1 Table. Ethics committees approving individual studies.** Approval for the analysis presented here was granted by the Observational / Interventions Research Ethics Committee of the London School of Hygiene and Tropical Medicine (ref #29931), and covers inclusion of data from the studies whose primary ethical approvals are listed in this table.

**S2 Table. Studies included in the analysis.** Details of type of study, number of isolates; laboratory methods for identification, storage, DNA extraction, sequencing; informatics methods for quality control and assembly; BioProject accession and citation/s for primary study.

**S3 Table. Isolates and sequence data included in the study.**

**S4 Table. Clusters identified in the study.**

**S5 Table. Summary of *Klebsiella pneumoniae* STs identified in the study.**

**S6 Table. Chi squared tests of resistance distribution by cluster status.** Only one isolate was included per cluster, and the representative cluster isolate was assigned the consensus ESBL or carbapenemase status for that cluster (e.g. if 3 of 4 cases in the cluster were ESBL, the cluster was recorded as ESBL-positive).

**S7 Table. Logistic regression model for transmission.** Multivariable logistic regression model to assess factors associated with transmission clusters. Each unique ‘introduction’ (i.e. cluster, or unclustered singleton case) was treated as an observation, with the outcome recorded as transmission (cluster size ≥2) or no transmission (unclustered singleton case). For each cluster, each predictor was assigned the consensus value for that cluster (e.g. if 3 of 4 cases were ESBL, the cluster was recorded as ESBL). Predictors ST, Site, K locus, and O type were encoded categorical variables. AMR variables (ESBL gene and carbapenemase gene presence) were encoded as binary. Commonly transmitted STs were defined as those that were identified in ≥3 transmission clusters in two or more sites; other STs were categorised as ‘Other’ and used as the reference category. The top 10 K loci were encoded as individual categories and compared to all other K loci as the reference category. O types were compared to the most common O type (51.6%) as the reference category. ESBL+: ESBL or carbapenemase gene carriage (Kleborate resistance score ≥ 1); CP+: carbapenemase gene carriage (Kleborate resistance score ≥ 2).

**S8 Table. Chi squared tests of resistance distribution by ST type.** Only one isolate was included per cluster, and the representative cluster isolate was assigned the consensus ESBL or carbapenemase status for that cluster (e.g. if 3 of 4 cases in the cluster were ESBL, the cluster was recorded as ESBL-positive).

**S9 Table. Summary of O serogroups detected.** Only one isolate was counted per cluster, and the representative cluster isolate was assigned the consensus ESBL or carbapenemase status for that cluster (e.g. if 3 of 4 cases in the cluster were ESBL, the cluster was recorded as ESBL-positive).

**S10 Table. Summary of K loci detected.** Only one isolate was counted per cluster, and the representative cluster isolate was assigned the consensus ESBL or carbapenemase status for that cluster (e.g. if 3 of 4 cases in the cluster were ESBL, the cluster was recorded as ESBL-positive).

**S1 Figure. Flow diagram for inclusion of isolates.** All numbers shown represent the number of isolates. For the line ‘Sites with N<10 pass filters’, the numbers represent the total number of isolates excluded for all excluded sites.

**S2 Figure. Geotemporal distribution of isolates included in the analysis.** (A) Location of study sites. Each point represents a unique site and point sizes indicate the number of isolates included per site, as per figure legend. Country colours indicate the number of isolates included per country, as per figure legend. The map was generated using the rnaturalearth (version 1.0.1) package in R. Base map source: Natural Earth, https://www.naturalearthdata.com/downloads/10m-cultural-vectors/, accessed using rnaturalearth R package v1.0.1, terms of use: https://www.naturalearthdata.com/about/terms-of-use/. (B) Plot shows sampling dates for all isolates included per sites. Each point represents a unique isolate included per site and numbers inside the box represent the number of isolates per site. Points are jittered along the y-axis to aid visibility.

**S3 Figure. Transmission clusters of *K. pneumoniae* neonatal sepsis cases per study.** Each point represents one or more cases isolated on specific dates. Points are coloured according to sequence type. Clusters are represented as groups of cases (points) linked by horizontal lines. Clusters belonging to the same sequence type are jittered along the y-axis to allow visibility of overlapping clusters. ST – sequence type. * – includes single locus variants of the respective STs.

**S4 Figure. Sensitivity of the cluster proportion estimates to varying temporal and genetic distance thresholds.** The sub-panels show estimates for individual study datasets at different combinations of genetic distance threshold (x-axis) and temporal distance threshold ranges (as per figure legend).

**S5 Figure. Cluster proportion vs facility characteristics.** (A) Individual site characteristics and estimates of proportion of cases in clusters. (B) Distribution of cluster proportion estimates by facility characteristics. Panels compare the distribution of proportion of isolates in clusters across different facility characteristics: (i) Onsite availability of neonatal surgical facilities (ii) Facility size (iii) Availability of piped water (iv) Number of neonatal beds. Individual points represent cluster proportions for specific facilities. Facility size – Small: 50–150 beds, Medium: 151–300 beds, Large: 301–600 beds, Very Large: >600 beds. CI: confidence interval

**S1 Appendix. Sensitivity of cluster analysis.**

**S1 Checklist. STrengthening the Reporting of OBservational studies in Epidemiology (STROBE) Statement – checklist of items that should be included in reports of observational studies, available at** https://www.strobe-statement.org/**, licenced under CC BY 4.0.**

## Author contribution statement

**Conceptualization:** Kathryn E Holt

**Data Curation:** Jabir A Abdulahi, Anne V Amulele, Matthew Bates, Eva Heinz, Weiming Hu, Kajal Jain, Rindidzani Magobo, Courtney P Olwagen, John M Tembo, Tolbert Sonda, Jonathan Strysko, Caroline C Tigoi

**Formal Analysis:** Erkison Ewomazino Odih, Kathryn E Holt

**Funding Acquisition:** Ramesh K Agarwal, Alexander M Aiken, James A Berkley, Susan E Coffin, Nicholas A Feasey, Nelesh P Govender, Davidson H Hamer, Kathryn E Holt, Shabir A Madhi, M Jeeva Sankar, Kelly L Wyres

**Investigation:** Kyle Bittinger, Jennifer Cornick, Ebenezer Foster-Nyarko, Wilson Gumbi, Steven M Jones, Chileshe Lukwesa, Carolyn M McGann, Ahmed M Moustafa, Patrick Musicha, James CL Mwansa, Moreka L Ndumba, Donwilliams O Omuoyo, Oliver Pearse, Laura T Phillips, Paul J Planet, Charlene MC Rodrigues, Fatou Secka, Kirsty Sands, Jonathan Strysko, Erin Theiller, Allan M Zuza

**Methodology:** Erkison Ewomazino Odih, Kathryn E Holt, Kelly L Wyres

**Resources:** Sulagna Basu, Grace J Chan, Kenneth C Iregbu, Jean-Baptiste Mazarati, Semaria Solomon Alemayehu, Timothy R Walsh, Rabaab Zahra, Kajal Jain, Angela Dramowski, Sombo Fwoloshi, Appiah-Korang Labi, Lola Madrid, Noah Obeng-Nkrumah, David Ojok, Boaz D Wadugu, Andrew C Whitelaw, Anudita Bhargava, Atul Jindal, Ramesh K Agarwal, M Jeeva Sankar, Alexander M Aiken, James A Berkley, Susan E Coffin, Nicholas A Feasey, Nelesh P Govender, Davidson H Hamer, Shabir A Madhi,

**Software:** Erkison Ewomazino Odih, Thomas D Stanton

**Supervision:** Sulagna Basu, Grace J Chan, Kenneth C Iregbu, Jean-Baptiste Mazarati, Semaria Solomon Alemayehu, Timothy R Walsh, Rabaab Zahra, Kajal Jain, Angela Dramowski, Sombo Fwoloshi, Appiah-Korang Labi, Lola Madrid, Noah Obeng-Nkrumah, David Ojok, Boaz D Wadugu, Andrew C Whitelaw, Anudita Bhargava, Atul Jindal, Ramesh K Agarwal, M Jeeva Sankar, Alexander M Aiken, James A Berkley, Susan E Coffin, Nicholas A Feasey, Nelesh P Govender, Davidson H Hamer, Shabir A Madhi,

**Visualization:** Erkison Ewomazino Odih, Kathryn E Holt

**Writing – Original Draft Preparation:** Erkison Ewomazino Odih, Kathryn E Holt

**Writing – Review & Editing:** All authors.

