## Supplementary material for "Contribution of nosocomial transmission to *Klebsiella pneumoniae* neonatal sepsis in Africa and South Asia: analysis of infection clusters inferred from pathogen genomics and temporal data": S1 Appendix

### S1 Appendix: Sensitivity of cluster analysis

#### Sensitivity to distance thresholds

Exploratory analyses of the sensitivity of clustering to choice of thresholds revealed that varying the SNV threshold within the range of 5 to 25 SNVs had minimal effect on the produced estimates (**Figures 1, 2**). Within this distance threshold range (5–25 SNVs), we observed a mean difference of 3.8% (range 0–12%) in cluster proportion estimates and a mean difference of 3.5% (range 0–11%) in transmission proportion estimates, calculated as the mean of the differences between the estimates at 5 vs 25 SNVs thresholds at a fixed temporal threshold (4 weeks) and across all datasets. Below 5 SNVs, cluster proportion estimates varied more substantially (mean difference 21.9%, range 10–43%, between 0 and 5 SNVs), with the greatest change observed from 0 to 1 SNVs (mean difference 11.7%, range 0–24%), and stabilising approaching 5 (mean difference 1.6%, range 0–6%, between 4 and 5 SNVs). The cluster and transmission proportion estimates remained stable beyond 25 SNVs, even up to an extreme SNV threshold value of 1,000 (**Figure 3**).

Compared to the genetic distance threshold, varying the temporal distance threshold between specimen collection dates had more of an impact on the transmission estimates. Nevertheless, there were minimal differences in the cluster proportion (mean difference =

7.3%, range 4–13%) and transmission proportion (mean difference = 7.3%, range 4–10%) estimated using 2 vs 8 weeks and a fixed genetic distance threshold of 10 SNVs. Further reducing the threshold to 1 week (i.e. only clustering isolates collected within 7 days) had a minimal impact in most sites (mean difference in cluster proportion of 4.4%, range 0–9%).

#### Sensitivity to bioinformatics approach to estimating genetic distance

To explore the sensitivity of our estimates to the choice of method for estimating genetic distance, we used a reference-free approach (SKA2) to estimate pairwise core-genome genetic distances for a subset of our *K. pneumoniae* data (isolates from BARNARDS [n=218], representing a multi-site study, and SPINZ [n=285], representing a large single-site study). This yielded very similar distance measures and near identical clustering results (see **Figure 4**).

#### Figure 1. Sensitivity of the cluster proportion estimates to varying temporal and genetic distance thresholds.

(reproduced from Figure S3 in main manuscript).

The sub-panels show estimates for individual study datasets at different combinations of genetic distance threshold (x-axis) and temporal distance threshold ranges (as per figure legend).

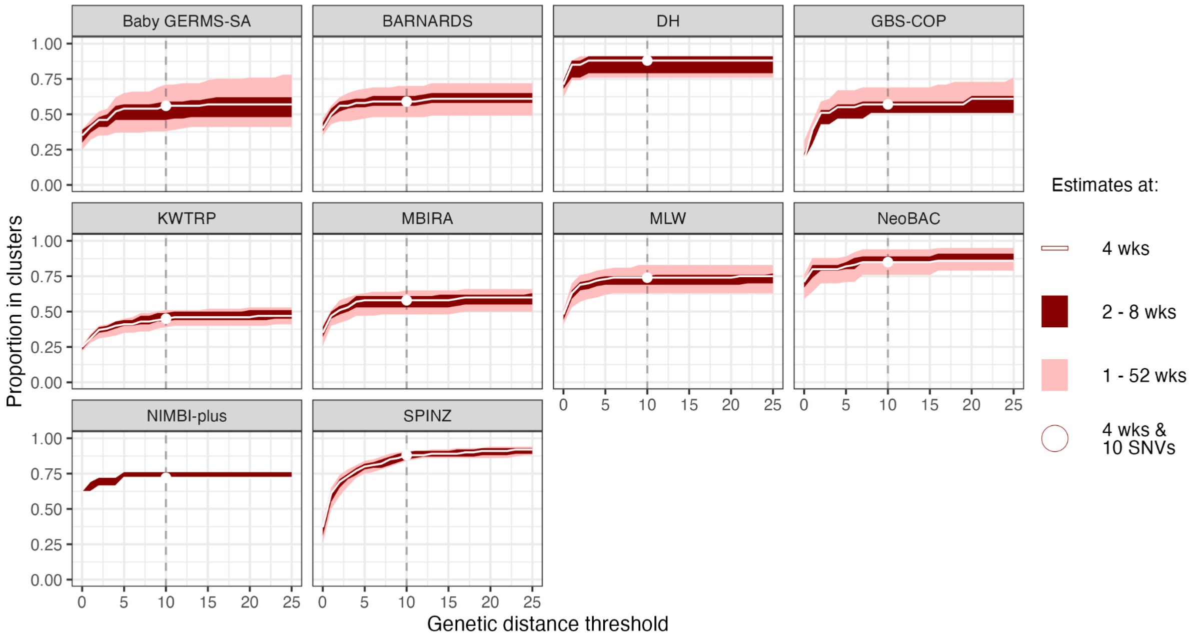

#### Figure 2. Sensitivity of the transmission proportion estimates to varying temporal and genetic distance thresholds.

The sub-panels show estimates for individual study datasets at different combinations of genetic distance threshold (x-axis) and temporal distance threshold ranges (as per figure legend).

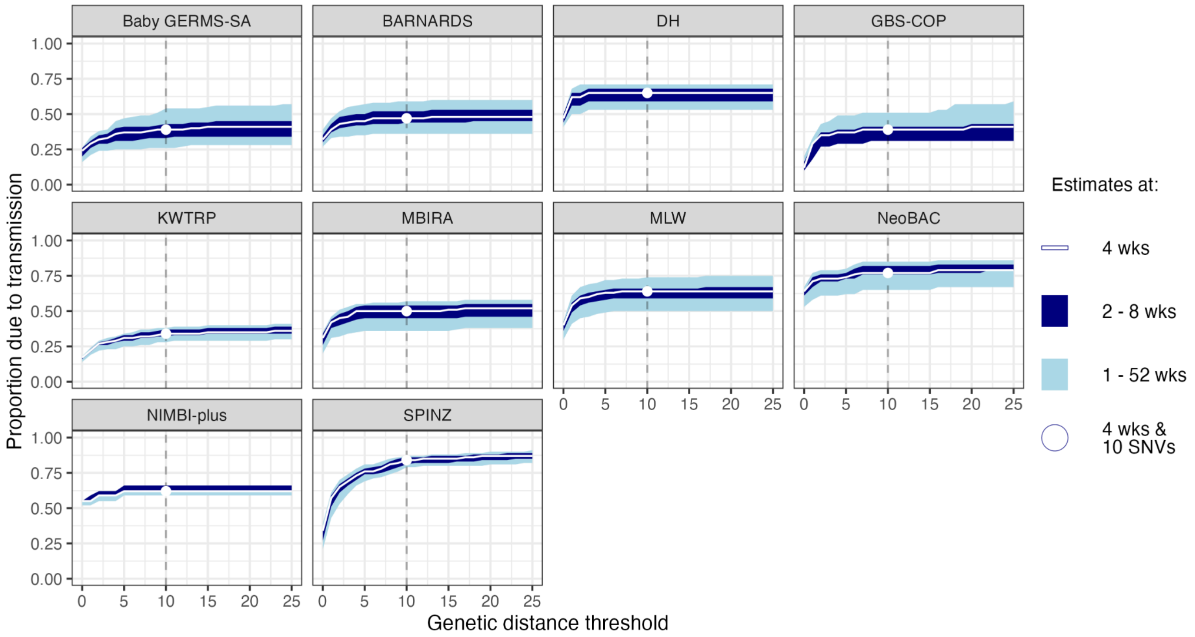

**Figure 3. Sensitivity of the cluster proportion and transmission proportion estimates to extreme genetic distance thresholds**

(A) Cluster proportion estimates (B) Transmission proportion estimates. The sub-panels show estimates across different temporal distance and extreme genetic distance threshold ranges, as per figure legend. SNP – single nucleotide polymorphism

**(A) Cluster proportion estimates**

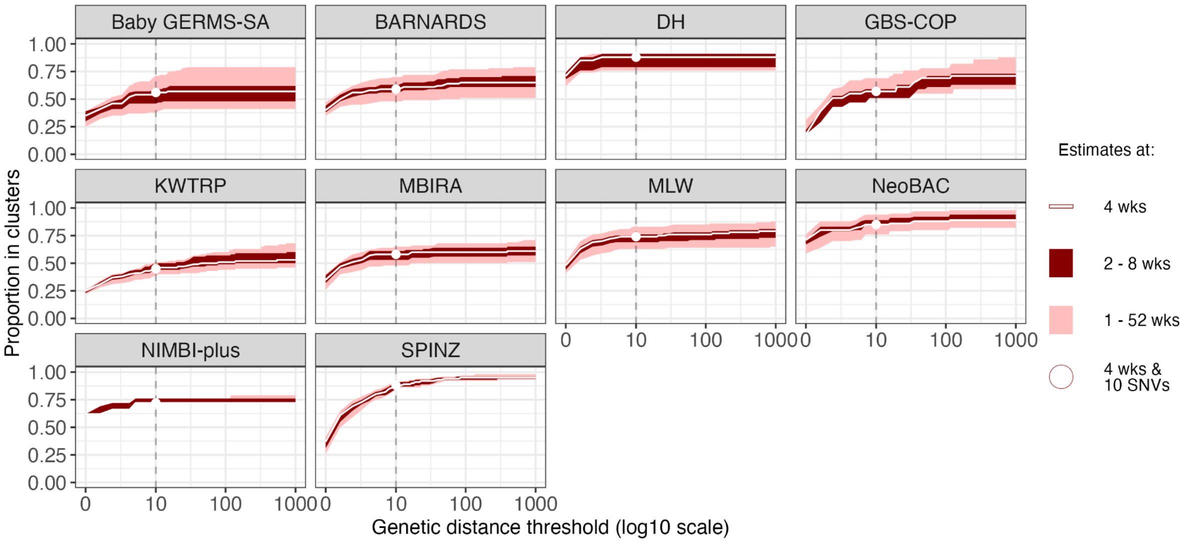


**(B) Transmission proportion estimates**

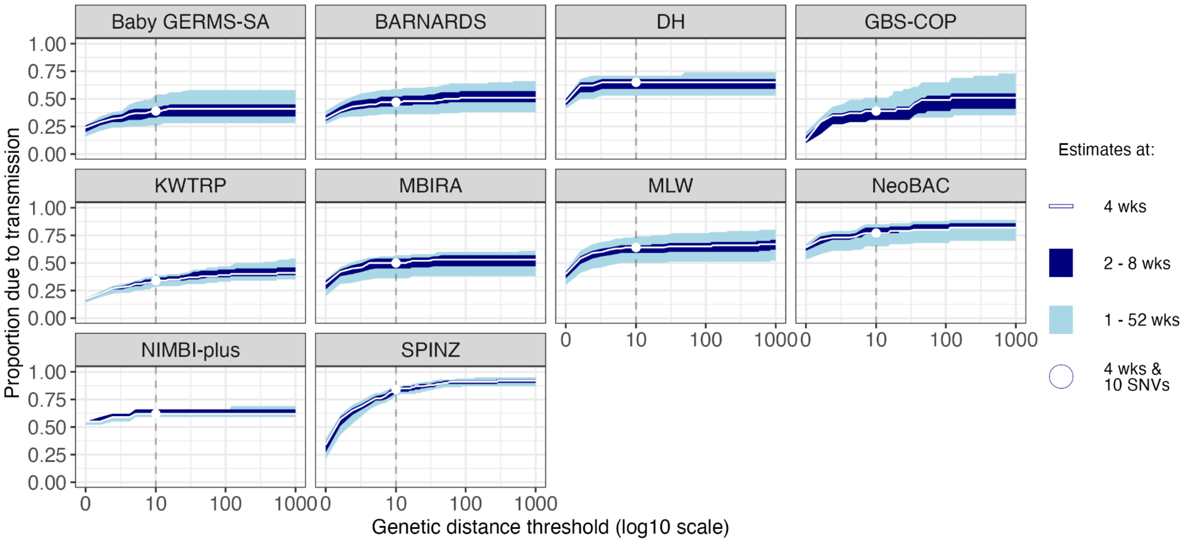

#### Figure 4. Impact of distance estimation method on transmission burden estimates

Plots show the cluster proportion (A) and transmission proportion (B) estimates across a range of genetic and temporal distance thresholds (as per figure legend), as well as the pairwise genetic distances estimated using Pathogenwatch and SKA for the (C) BARNARDS and (D) SPINZ datasets. Inset plots for (C) and (D) show pairwise genetic distances ≤100 SNVs. For the SPINZ dataset, at a distance threshold of 10 SNVs, the estimated cluster proportion at 4 weeks threshold was 0.88 (2-8 weeks threshold range: 0.85–0.90) using the Pathogenwatch distances compared to 0.86 (0.84-0.88) using the SKA distances. Across all sites in the BARNARDS dataset, identical cluster proportion (0.59 [0.56-0.63] vs 0.57 [0.53-0.61]) and transmission proportion (0.47 [0.44-0.52] vs 0.44 [0.41-0.50]) estimates were obtained using both the Pathogenwatch and SKA distances with a genetic distance threshold of 10 SNVs. PW – Pathogenwatch; SKA – split k-mer analysis; SNV – single nucleotide variant.

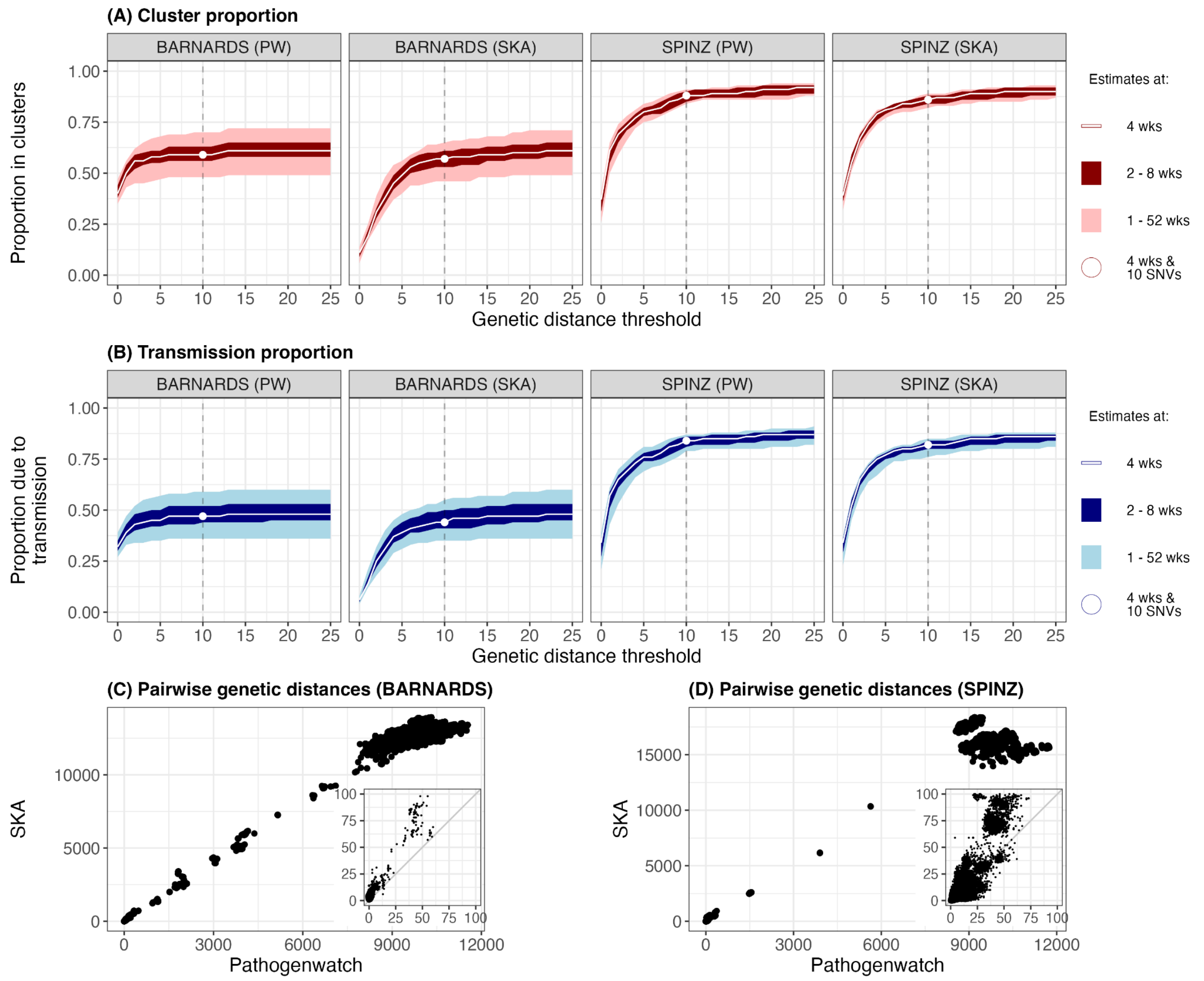
