## Supplemental Information for "Contribution of nosocomial transmission to *Klebsiella pneumoniae* neonatal sepsis in Africa and South Asia: analysis of infection clusters inferred from pathogen genomics and temporal data"

#### Code and data availability

All input data and code required to reproduce the results, figures, and tables presented in this paper are available at https://github.com/klebgenomics/KlebNNS_transmission (DOI: 10.5281/zenodo.17591910). The methods for clustering infections, quantifying transmission, and exploring the sensitivity of the estimates to selected thresholds (as described above) are implemented in a species-agnostic Shiny web application (https://klebsiella.shinyapps.io/transmission_estimator/), with source code available at https://github.com/klebgenomics/transmission_estimator (DOI: 10.5281/zenodo.17593948).

#### Supporting Tables

##### S1 Table. Ethics committees approving individual studies

Approval for the analysis presented here was granted by the Observational / Interventions Research Ethics Committee of the London School of Hygiene and Tropical Medicine (ref #29931), and covers inclusion of data from the studies whose primary ethical approvals are listed in this table.

| Study | Ethics Committees granting approval |
|---|---|
| **Baby GERMS-SA** | Human Research Ethics Committee of the University of the Witwatersrand (M190320). Approvals for the tier 2 surveillance study were received from each provincial research committee through registration on the National Health Research Database. |
| **BARNARDS** | Ethical Review Committee, Bangladesh Institute of Child Health, BICH-ERC-4/3/2015, 15/09/2015. Boston Children's Hospital, IRB-P00023058, 11/08/2016. Institutional Ethics Committee, National Institute of Cholera and Enteric Diseases and Institute of Post Graduate Medical Education and Research, A-I/2016-IEC, 17/11/2016. IPGMER Research Oversight Committee, Inst/IEC/2016/508, 04/11/2016. Kano State Hospitals Management Board, 8/10/1437AH, 13/07/2016. Health Research Ethics Committee (HREC), National Hospital, Abuja, NHA/EC/017/2015, 27/04/2015. Republic of Rwanda National Ethics Committee, No342/RNEC/2015, 10/11/2015. Stellenbosch University and Tygerberg Hospital, Research projects, Western Cape Government, N15/07/063, 04/12/2015 and 02/02/2016. |
| **KWTRP surveillance** | KEMRI Scientific and Ethics Review Unit, ref 281/4687, 17/4/2023. A research license was obtained from the National Commission for Science, Technology and Innovation, ref 527823, license no. NACOSTI/P/23/26005, 31/5/2023. |
| **GBS-COP** | University of the Witwatersrand, Human Research Ethics Committee (HREC), ref 181110, 24/4/2023. |
| **MLW Biobank** | University of Malawi College of Medicine Research Ethics Committee (COMREC) (P.11/18/2541). |
| **SPINZ** | Boston University Medical Center Institutional Review Board, USA (ref H-33473), 21/3/2016. Excellence in Research Ethics and Science (ERES) CONVERGE, Zambia (ref 2015-Jan-004), 1/3/2015. |
| **NIMBIplus** | NIMBI study: University of Pennsylvania IRB (ref 833786), 22/4/2020. Children’s Hospital of Philadelphia Research Institute IRB (ref 19-016848), 25/7/2020. IRBs of Princess Marina Hospital IRB (PMH 2/11AII(372)), 20/3/2024 and the Health Research Development Committee (HRDC) in Botswana (ref HPRD 6/14/1), 15/2/2024. |

| | |
|---|---|
| | SHARE study: University of Pennsylvania IRB (ref 851492), 9/6/2020. Princess Marina Hospital (ref PMH 2/2A(7)/201), 20/5/2022. University of Botswana IRB (ref UBR/RES/IRB/BIO/205), 11/5/2022. Health Research & Development Committee (HRDC) in Botswana (HPDME:13/18/1), 21/4/2022. |
| **MBIRA** | London School of Hygiene and Tropical Medicine HREC, ref 21236-4, latest amendment approved 27/3/2024. Korle Bu Teaching Hospital Institutional Review Board, 00097/2020, 18/11/2020; National Health Research Authority, 17/2/2020; Ministry of Science and Higher Education, 1.16/10.81/13, 24/2/2021; Western Cape Government, N20/02/072; National Institute for Medical Research, R.8a/Vol.IX/3575, 11/12/2020; Combined Research and Ethics Committee Swarm Hub research protocol v2, 2/11/2020. |
| **DH** | Institute Ethics Committee, All India Institute of Medical Sciences, New Delhi, ref IEC-683/07.12.2018, RP-12/2018, 18/12/2018. |

#### S2 Table. Studies included in the analysis.

Details of type of study, number of isolates; laboratory methods for identification, storage, DNA extraction, sequencing; informatics methods for quality control and assembly; BioProject accession and citation/s for primary study.
https://github.com/klebgenomics/KlebNNS_transmission/blob/main/tables/TableS2_StudyMethodsInfo.tsv

#### S3 Table. Isolates and sequence data included in the study

https://github.com/klebgenomics/KlebNNS_transmission/blob/main/tables/TableS3_SampleInfo.tsv

#### S4 Table. Clusters identified in the study

https://github.com/klebgenomics/KlebNNS_transmission/blob/main/tables/TableS4_ClustersInfo.tsv

#### S5 Table. Summary of *Klebsiella pneumoniae* STs identified in the study

https://github.com/klebgenomics/KlebNNS_transmission/blob/main/tables/TableS5_ClonesSummary.tsv

#### S6 Table. Chi squared tests of resistance distribution by cluster status

Only one isolate was included per cluster, and the representative cluster isolate was assigned the consensus ESBL or carbapenemase status for that cluster (e.g. if 3 of 4 cases in the cluster were ESBL, the cluster was recorded as ESBL-positive).

| **Test** | **Category** | **Clusters** | **Singletons** | ***P*** |
|---|---|---|---|---|
| ESBL gene presence | No | 9 (5.8%) | 89 (18.2%) | **0.00027** |
| | Yes | 147 (94.2%) | 399 (81.8%) | |
| Carbapenemase gene presence | No | 125 (80.1%) | 438 (89.8%) | **0.0025** |
| | Yes | 31 (19.9%) | 50 (10.2%) | |

#### S7 Table. Logistic regression model for transmission

Multivariable logistic regression models to assess factors associated with transmission clusters. Each unique 'introduction' (i.e. cluster, or unclustered singleton case) was treated as an observation, with the outcome recorded as transmission (cluster size ≥2) or no transmission (unclustered singleton case). For each cluster, each predictor was assigned the consensus value for that cluster (e.g. if 3 of 4 cases were ESBL, the cluster was recorded as ESBL). Predictors ST, Site, K locus, and O type were encoded categorical variables. AMR variables (ESBL and carbapenemase gene presence) were encoded as binary. Commonly transmitted STs were defined as those that were identified in ≥3 transmission clusters in two or more sites; other STs were categorised as 'Other' and used as the reference category. The top 10 K loci were encoded as individual categories and compared to all other K loci as the reference category. O types were compared to the most common O type (51.6%) as the reference category. ESBL+: ESBL or carbapenemase gene carriage (Kleborate resistance score ≥ 1); CP+: carbapenemase gene carriage (Kleborate resistance score ≥ 2).

https://github.com/klebgenomics/KlebNNS_transmission/blob/main/tables/TableS7_TransmissionPredictors.tsv

#### S8 Table. Chi squared tests of resistance distribution by ST type

Only one isolate was included per cluster, and the representative cluster isolate was assigned the consensus ESBL or carbapenemase status for that cluster (e.g. if 3 of 4 cases in the cluster were ESBL, the cluster was recorded as ESBL-positive).

| **Test** | **Category** | **Most commonly transmitted STs (≥3 clusters in ≥2 sites)** | **Other STs** | ***P*** |
|---|---|---|---|---|
| ESBL gene presence | No | 19 (5.8%) | 79 (24.9%) | **3.13E-11** |
| | Yes | 308 (94.2%) | 238 (75.1%) | |
| Carbapenemase gene presence | No | 281 (85.9%) | 282 (89.0%) | 0.30 |
| | Yes | 46 (14.1%) | 35 (11.0%) | |

#### S9 Table. Summary of O serogroups detected

Only one isolate was counted per cluster, and the representative cluster isolate was assigned the consensus ESBL or carbapenemase status for that cluster (e.g. if 3 of 4 cases in the cluster were ESBL, the cluster was recorded as ESBL-positive).

| O group | No. isolates | | | No. countries | | | No. sites | | | No. STs | | |
|---|---|---|---|---|---|---|---|---|---|---|---|---|
| | All | Clusters | Singletons | All | Clusters | Singletons | All | Clusters | Singletons | All | Clusters | Singletons |
| **O1** | 332 (51.6%) | 80 (51.3%) | 252 (51.6%) | 13 | 13 | 13 | 27 | 22 | 27 | 78 | 29 | 76 |
| **O2** | 158 (24.5%) | 42 (26.9%) | 116 (23.8%) | 13 | 9 | 13 | 25 | 15 | 24 | 46 | 15 | 43 |
| **O3** | 52 (8.1%) | 6 (3.8%) | 46 (9.4%) | 12 | 4 | 12 | 18 | 5 | 16 | 34 | 5 | 33 |
| **O4** | 41 (6.4%) | 13 (8.3%) | 28 (5.7%) | 9 | 6 | 8 | 15 | 9 | 11 | 12 | 8 | 11 |
| **O5** | 35 (5.4%) | 10 (6.4%) | 25 (5.1%) | 5 | 4 | 5 | 12 | 8 | 9 | 4 | 1 | 4 |
| **O13** | 22 (3.4%) | 4 (2.6%) | 18 (3.7%) | 6 | 3 | 6 | 7 | 3 | 7 | 14 | 3 | 12 |
| **unknown** | 4 (0.6%) | 1 (0.6%) | 3 (0.6%) | 4 | 1 | 3 | 4 | 1 | 3 | 4 | 1 | 3 |

#### S10 Table. Summary of K loci detected

Only one isolate was counted per cluster, and the representative cluster isolate was assigned the consensus ESBL or carbapenemase status for that cluster (e.g. if 3 of 4 cases in the cluster were ESBL, the cluster was recorded as ESBL-positive).
https://github.com/klebgenomics/KlebNNS_transmission/blob/main/tables/TableS10_KLSummary.tsv

### Supporting Figures

#### S1 Figure. Flow diagram for inclusion of isolates

All numbers shown represent the number of isolates. For the line ‘Sites with N<10 pass filters’, the numbers represent the total number of isolates excluded for all excluded sites.

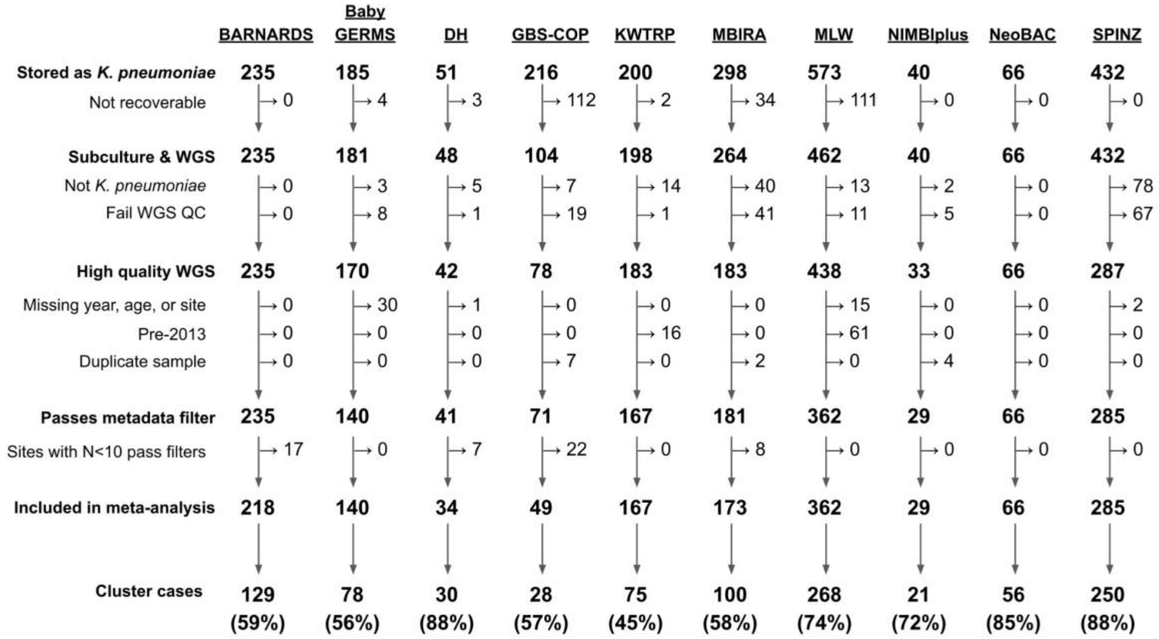

S2 Figure. Geotemporal distribution of isolates included in the analysis

(A) Location of study sites. Each point represents a unique site and point sizes indicate the number of isolates included per site, as per figure legend. Country colours indicate the number of isolates included per country, as per figure legend. The map was generated using the rnaturalearth (version 1.0.1) package in R. Base map source: Natural Earth, https://www.naturalearthdata.com/downloads/10m-cultural-vectors/, accessed using rnaturalearth R package v1.0.1, terms of use: https://www.naturalearthdata.com/about/terms-of-use/. (B) Plot shows sampling dates for all isolates included per sites. Each point represents a unique isolate included per site and numbers inside the box represent the number of isolates per site. Points are jittered along the y-axis to aid visibility.

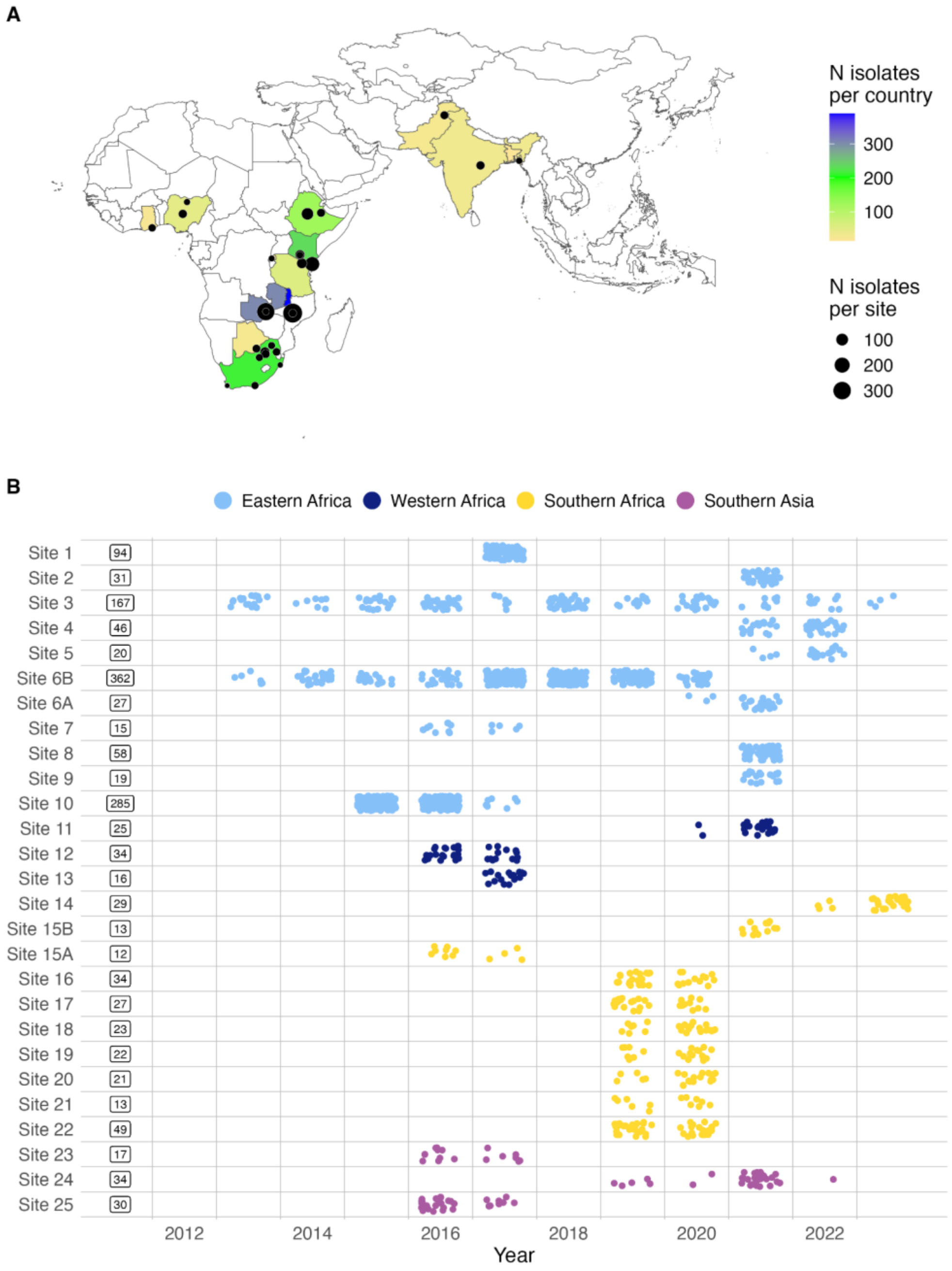

#### S3 Figure. Transmission clusters of *K. pneumoniae* neonatal sepsis cases per study

Each point represents one or more cases isolated on specific dates. Points are coloured according to sequence type. Clusters are represented as groups of cases (points) linked by horizontal lines. Clusters belonging to the same sequence type are jittered along the y-axis to allow visibility of overlapping clusters. ST – sequence type. * – includes single locus variants of the respective STs.

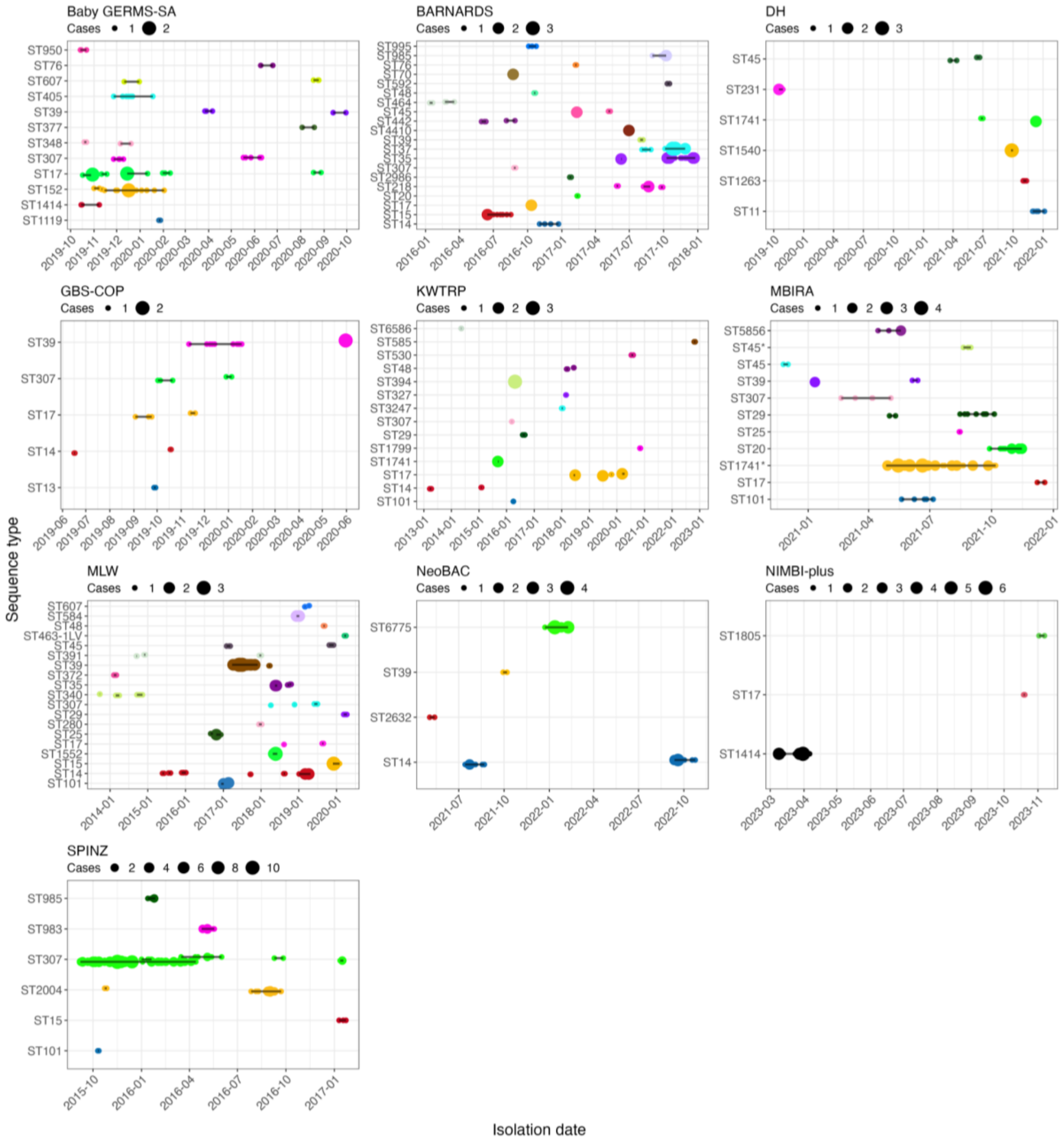

S4 Figure. Sensitivity of the cluster proportion estimates to varying temporal and genetic distance thresholds.

The sub-panels show estimates for individual study datasets at different combinations of genetic distance threshold (x-axis) and temporal distance threshold ranges (as per figure legend).

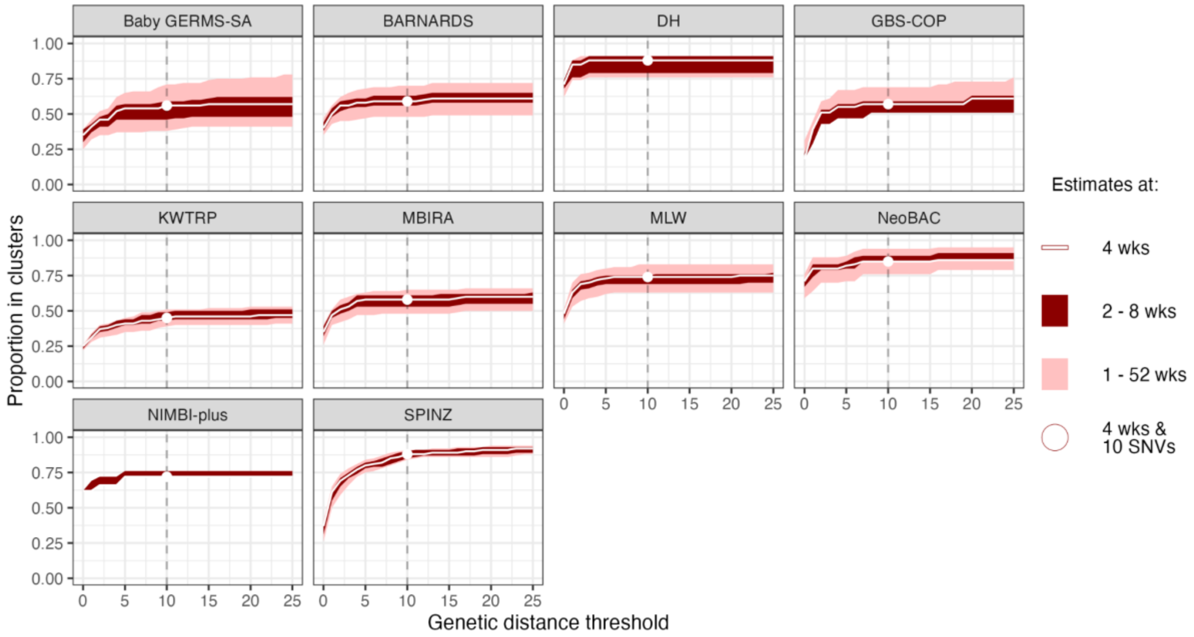

#### S5 Figure. Cluster proportion vs facility characteristics

(A) Individual site characteristics and estimates of proportion of cases in clusters. (B) Distribution of cluster proportion estimates by facility characteristics. Panels compare the distribution of proportion of isolates in clusters across different facility characteristics: (i) Onsite availability of neonatal surgical facilities (ii) Facility size (iii) Availability of piped water (iv) Number of neonatal beds. Individual points represent cluster proportions for specific facilities. Facility size – Small: 50–150 beds, Medium: 151–300 beds, Large: 301–600 beds, Very Large: >600 beds. CI: confidence interval

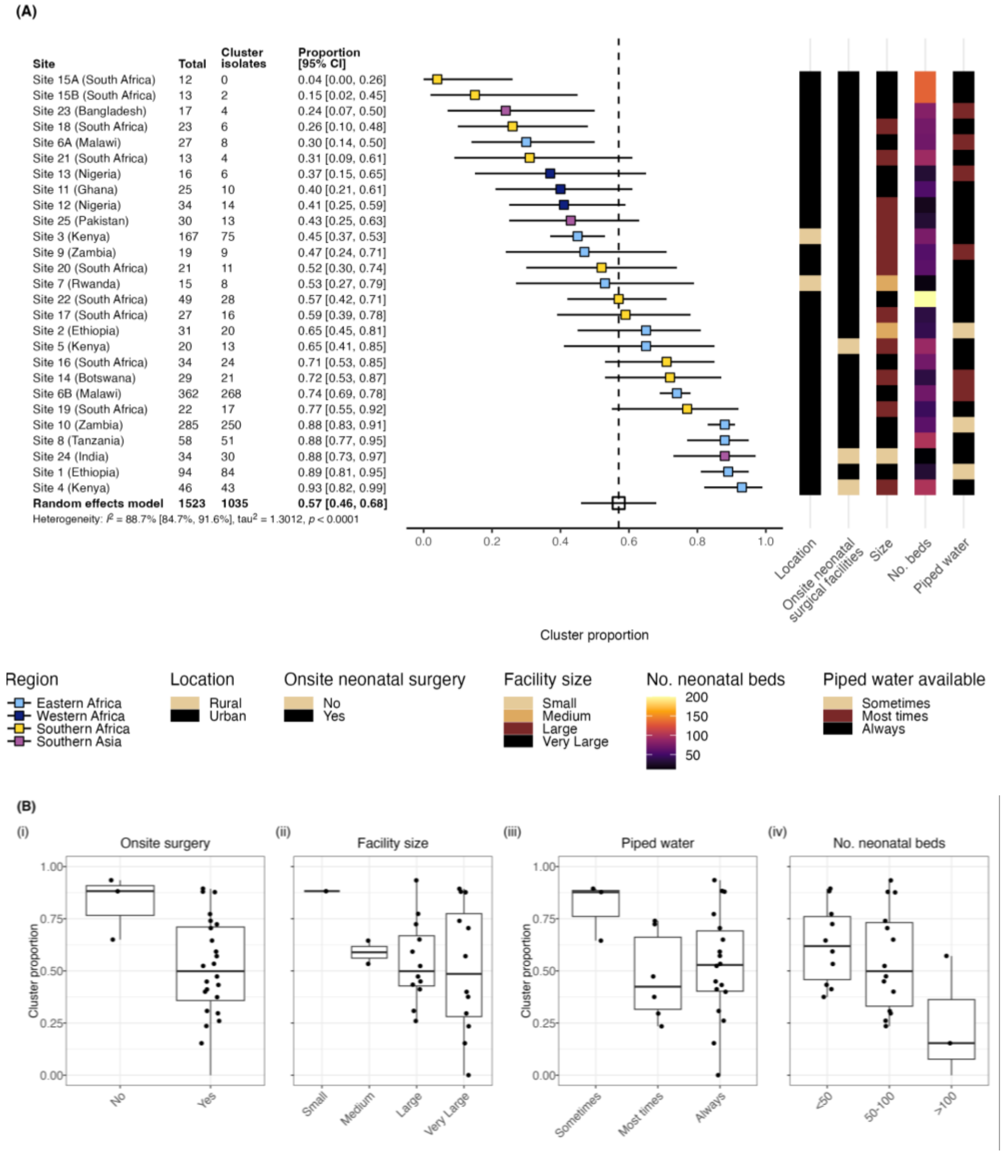
